# Transdiagnostic Psychiatric Polygenic Scores Reveal Pleiotropic Effects Across Medical and Laboratory Phenome

**DOI:** 10.64898/2026.11.10.26367763

**Authors:** Yingjie Shi, Maria Niarchou, Yingzhe Zhang, Younga Heather Lee, Karmel W. Choi, Freida Blostein, Peter Straub, Tian Ge, Yen-Chen Anne Feng, Nina Roth Mota, Emma Sprooten, Barbara Franke, Lea K. Davis

**Author notes:** Corresponding author: Barbara Franke Route 200, Kapittelweg 29 6525 EN Nijmegen, The Netherlands. Equal contribution (Maria Niarchou, Yingzhe Zhang, Younga Heather Lee, Barbara Franke, Lea K. Davis).

## Abstract

**Background:** Current psychiatric diagnoses lack biological grounding. Using transdiagnostic psychiatric dimensions derived through data-driven genetic studies with large-scale clinical health records of multiple physiological and clinical outcomes, we can gain new insights into biological similarity and differences between such dimensions, informing nosological discourse.

**Methods:** Here, we examined data from 108,182 individuals of European-like ancestry and 13,229 African-like ancestry across two healthcare biobanks. Genetic liabilities of individuals were estimated as polygenic scores for five first-order psychopathology factors (compulsive, thought, neurodevelopmental, substance use, internalizing disorders) and one second-order factor (hierarchical/p-factor). Phenome-wide and laboratory-wide association scans encompassed 1,520 medical phenotypes and 123 laboratory metrics, meta-analyzed across sites. People with lived experience were involved in the design, preparation, interpretation and reporting of this work.

**Outcomes:** We observed robust associations with up to 143 conditions (14 disease categories) for first- order factors and 221 conditions (15 categories) for the hierarchical factor (OR=0.75-1.94). Circulatory, digestive, and respiratory diseases were linked to neurodevelopmental, internalizing, and substance use liabilities. Metabolic conditions were negatively associated with thought and compulsive liabilities but positively associated with the remaining three factors. Laboratory results, particularly lipid and glucose metabolism markers, supported these differential findings. Immune markers also showed differential associations, with C- reactive protein and Complement C4 specifically linked to neurodevelopmental and thought factors, respectively. Similar trends emerged in the smaller African-like ancestry sample.

**Interpretation:** Overall, these findings support ongoing efforts to define biologically informed subtypes in psychiatry, such as immuno-metabolic profiles, which can facilitate patient stratification in clinical trials and support transdiagnostic precision psychiatry approaches.

**Funding:** The work was supported by funding from the participating institutions. BF’s and NRM’s contributions were supported by the European Community’s Horizon 2020 Programme (H2020/2014–2020) under grant agreement No. 847879 (PRIME). NRM was also supported by the Netherlands Organisation for Scientific Research (NWO) Innovation Programme (Vidi grant No. 09150172410043). Research reported in this publication was also supported by the National Institute of Mental Health of the National Institutes of Health under Award Number R01MH124851. The content is solely the responsibility of the authors and does not necessarily represent the official views of the funders.

**Research in Context:** 

**Evidence before this study:** We searched PubMed from database inception to October 13, 2026, without date or language restrictions, using combinations of the terms “transdiagnostic”, “cross-disorder”, “genomic SEM”, “polygenic score”, “polygenic risk”, “phenome-wide”, “PheWAS”, “laboratory”, and “somatic”. We considered original human genetic studies examining transdiagnostic psychiatric genetic liability and its associations with medical or laboratory phenotypes, excluding studies without relevant psychiatric genetic analyses. Previous studies demonstrated shared genetic liability across psychiatric disorders and phenome-wide associations of cross-disorder polygenic scores, although evidence was limited by ancestry representation, sample selection, and phenotypic ascertainment. Studies jointly examining multiple transdiagnostic psychiatric genetic dimensions across medical diagnoses and laboratory biomarkers remain limited.

**Added value of this study:** Using EHR-linked data from 121,411 individuals across two US biobanks, we examined six genetically defined transdiagnostic psychiatric factors in relation to 1,520 medical phenotypes and 123 laboratory measures. We identified shared and distinct associations across psychiatric dimensions, particularly involving metabolic, immune, and cardiovascular systems. Findings showed broadly concordant directions of association across genetically inferred ancestry groups, although statistical power was limited in the smaller African-like ancestry cohort.

**Implications of all the available evidence:** Our findings highlight the value of transdiagnostic genetic approaches for understanding shared and distinct physiological correlates of psychiatric liability beyond conventional diagnostic boundaries. These associations may inform future research into biologically informed psychiatric stratification and the integration of physical health considerations into psychiatric care, although their clinical utility requires further evaluation.

## Introduction

Psychiatric disorders are highly common, debilitating conditions with complex clinical presentations. Comorbidity with other psychiatric disorders and somatic conditions is high.^1^ Meanwhile, individuals with the same diagnosis exhibit significant phenotypic heterogeneity, while those with different diagnoses can display overlapping symptoms. Such clinical overlap and heterogeneity hinder etiological investigations under the conventional diagnostic classification framework, and have encouraged the discovery of underlying latent factors that transcend diagnostic entities.^2,3^

Joint modeling of the genetic architecture across various psychiatric disorders has revealed pervasive genetic overlap across diagnostic boundaries.^4^ Advanced statistical tools like genomic structural equation modeling (gSEM)^4^ have leveraged summary data from genome- wide association studies (GWAS) to pinpoint genetic loci associated with transdiagnostic psychopathology factors. Grotzinger et al. (2023)^5^ identified five transdiagnostic dimensions of genetic risk (i.e., compulsive, thought, neurodevelopmental, internalizing, and substance use disorders factors) along with a general psychopathology factor (i.e., hierarchical factor) that captures the shared variance among these dimensions. Aggregating the small effects of transdiagnostic dimension-associated alleles across the genome for each individual, polygenic scores (PGS)^5^ can provide a personal composite index for these latent dimensions of psychiatric risk. This can enhance the detection of associations between genetic susceptibility for psychiatric problems and downstream outcomes, promoting a deeper understanding of the biological processes underlying shared genetic risks across psychopathology.

Large-scale biobanks have linked electronic health records (EHRs) from clinical routine care with individuals’ genomic data. This enables the interrogation of the implications of genetic risk factors for the medical and biological phenome. The comprehensive, curated phenotypes in the EHRs, including diagnostic and procedural codes, clinical laboratory codes, and medication prescriptions, have been utilized in hypothesis-free designs with high-dimensional association analyses such as phenome-wide and laboratory-wide association scans (PheWAS^6,7^ and LabWAS^8^) to query unknown and novel gene-disease and gene-biomarker associations.

In this study, we leveraged comprehensive biobank data from two healthcare systems within the PsycheMERGE Network to explore the impact of polygenic risks derived from transdiagnostic psychopathology factors on diverse clinical markers and outcomes. Our goal was to elucidate the relationships between transdiagnostic psychiatric genetic liabilities and both diagnostic and laboratory markers in real-world settings. PGSs capturing genetic variations for six well-defined psychiatric dimensions were tested against the clinical phenome. A series of conditional PheWAS and LabWAS analyses were performed to parse the effects attributable to pleiotropy versus secondary risk factors. This data-driven approach allowed us to identify converging and distinct physiological correlates of biologically defined psychiatric dimensions, enhancing our understanding of their transdiagnostic etiology and providing novel insights into the interpretation of the co-occurrence of mental and physical conditions.

## Methods

### Study samples

This study used data generated from two biobanks within the PsycheMERGE network. The BioVU biobank consists of individuals who receive care at Vanderbilt University Medical Center (VUMC) and are willing to participate in the BioVU research study. The VUMC Institutional Review Board approved this project (IRB#172020). The participants in the Mass General Brigham Biobank (MGBB) were recruited during an in-person interaction with clinical research staff, through an electronic consent workflow, or through collaborating studies. The study was approved by the Institutional Review Board (or Ethics Committee) of Mass General Brigham (protocol code 2009-P-002312). Informed consent was obtained from all patients. In both biobanks, the genomic sample of each participant can be linked with extensive electronic health record data and clinical laboratory test results.

### Involvement of people with lived experience

People with lived experience were involved in the design, preparation, interpretation and reporting of this work.

### Statistical analyses

#### Polygenic scoring of the psychiatric genomic factors

To capture the joint genetic architecture across major psychiatric disorders, we adopted two multivariate genomic factor structures based on results from Grotzinger et al. (2023)^4^: the correlated factors model and the hierarchical factor model **(eMethods, eFigure 1**). The correlated factors model characterized five correlated first-order psychopathology factors based on genetic covariances among 13 major psychiatric disorders (see **eTable 1** for all contributing GWAS sources) and provided a good fit (CFI=0.95, SRMSR=0.102). These factors are: **compulsive disorders factor** (n_effective_=17,843) comprising anorexia nervosa (AN), obsessive-compulsive disorder (OCD), and Tourette syndrome (TS); **thought disorders factor** (n_effective_=127,485) comprising schizophrenia (SCZ) and bipolar disorder (BIP); **neurodevelopmental disorders factor** (n_effective_=37,492) comprising TS, autism spectrum disorder (ASD), attention-deficit/hyperactivity disorder (ADHD), post-traumatic stress disorder (PTSD), and major depressive disorder (MDD); **internalizing disorders factor** (n_effective_=370,650) comprising PTSD, MDD, and anxiety (ANX); **substance use disorders factor (SUD factor**, n_effective_=39,696) comprising alcohol use disorder (AUD), cannabis use disorder (CUD), and opioid use disorder (OUD). The hierarchical model captured the correlations among the five first-order factors with a second-order general psychopathology factor (hereafter referred to as the hierarchical factor, n_effective_=737,456) and provided a good fit (CFI=0.900; SRMSR=0.129). More information on the model estimation is described in **eMethods**.

**Figure 1.**
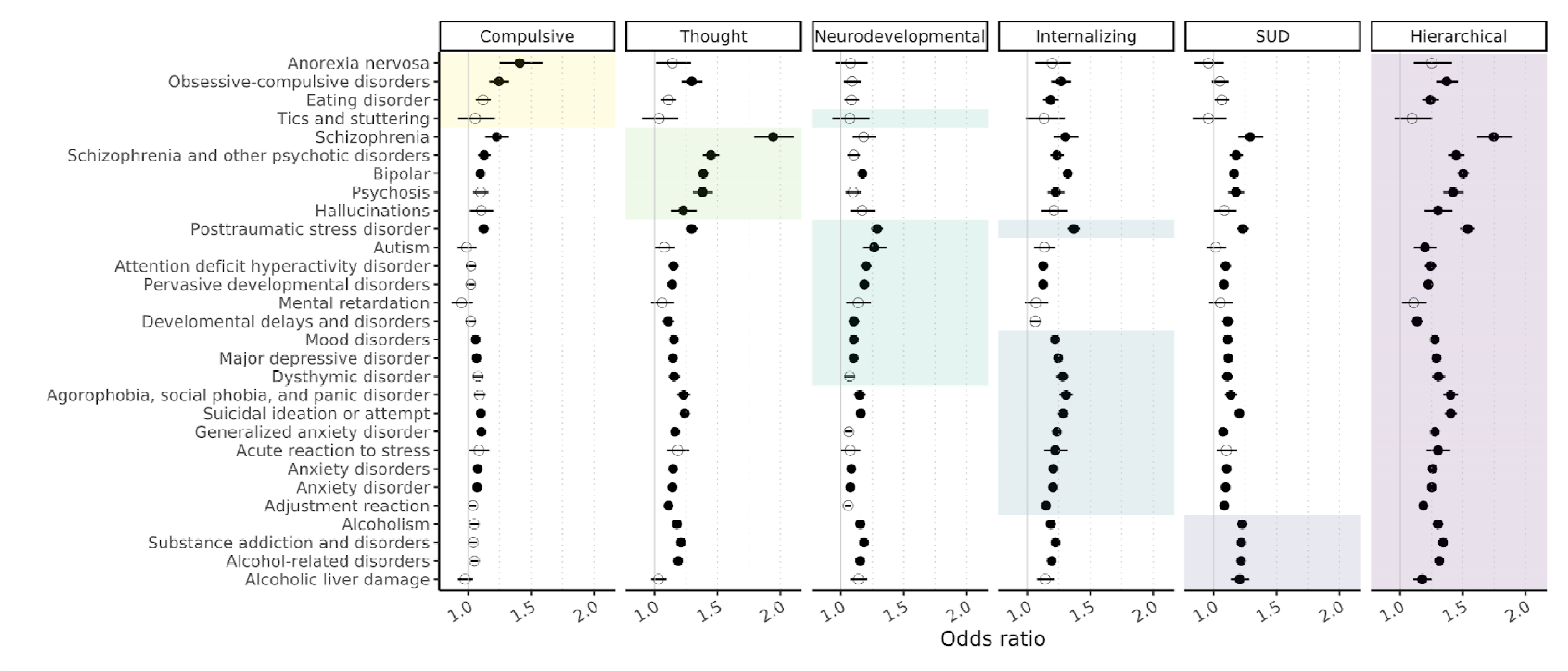
Meta-analyzed PheWAS of PGS for psychiatric genomic factors with mental disorders. The primary or closely related phenotypes used to esti psychiatric genomic factor structure are highlighted with background tiles corresponding to each factor. Filled dots indicate phenome-wide significant associat (α=0.05/(1520*6)=5.48e-6). The line segment indicates 95% confidence interval. SUD: substance use disorders.

**Figure 2.**
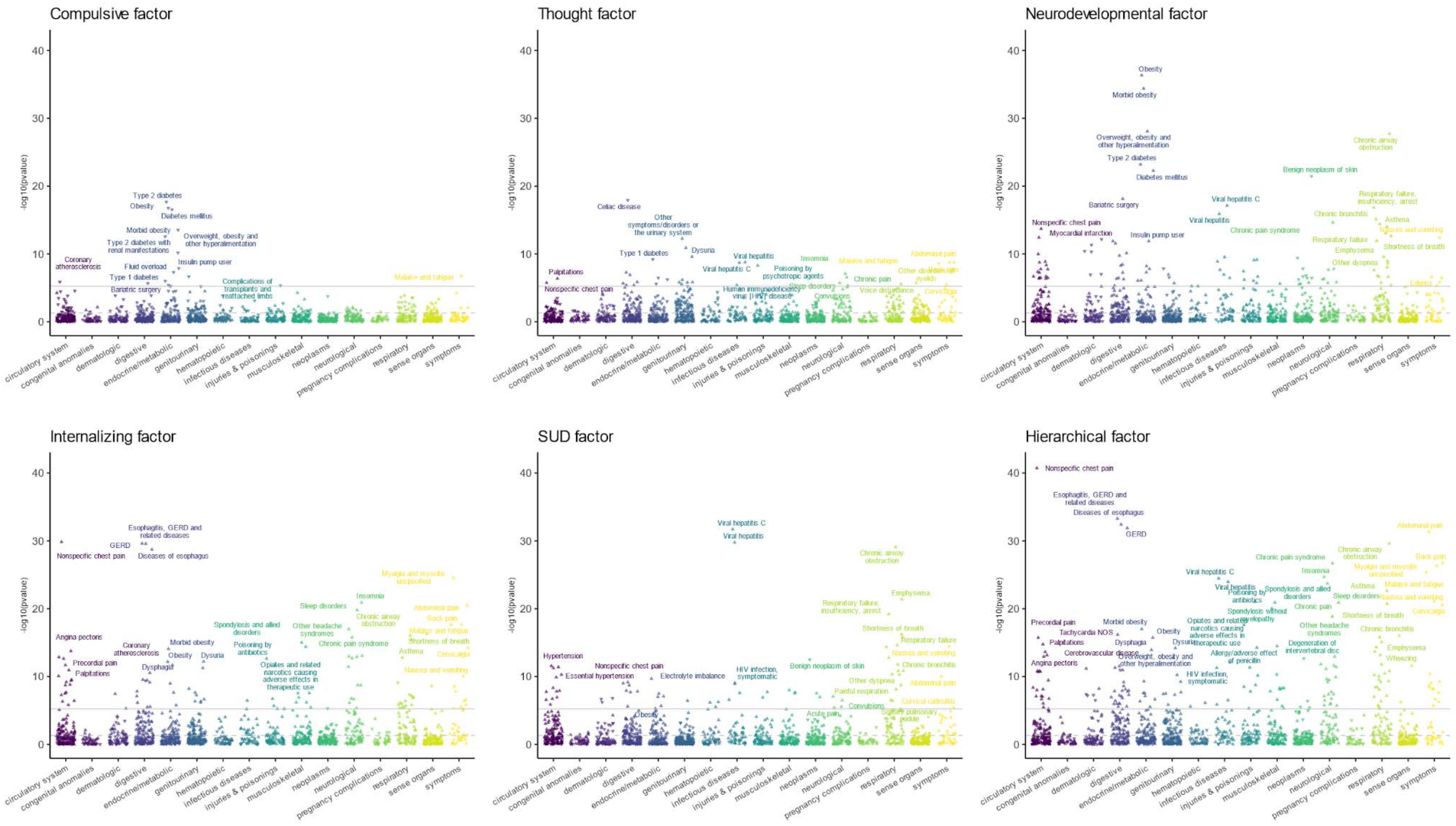
Meta-analyzed Phenome-wide association scan (PheWAS) of PGS for psychiatric genomic factors with all somatic diseases. Phenotypes are grouped by disease category on the x-axis; the significance of each PGS-phecode association is shown on the y-axis as -log_10_(p). The direction of the triangle indicates the direction of effect. The grey solid line indicates the Bonferroni-corrected significance threshold α=0.05/(1520*6)=5.48e-6, and the grey dotted line indicates α=0.05.

For each biobank, using the genotyped data after stringent quality control, we derived polygenic scores for the first- and second-order factors for each individual of genetically inferred European-like or African-like ancestry (**eMethods**). Polygenic scores were constructed using PRS-CS,^9^ which utilizes a Bayesian-based framework to infer single nucleotide polymorphism (SNP) effect sizes by applying continuous shrinkage.

#### PheWAS for genomic factor PGSs

To examine the clinical manifestations of transdiagnostic genetic liability captured by the multivariate genomic model, we conducted PheWAS using PGS of the genomic factors across the medical phenome, including 17 disease categories. Medical phenotypes (i.e., phecodes) tested were binary values generated from hierarchically clustered ICD-9 and ICD- 10 codes.^10,11^ At least two ICD codes on two separate days were required for an individual to be classified as a case for each phecode. Controls had no instances of qualifying case- defining codes and no additional exclusions were applied. The *PheWAS* R package was used to perform logistic regression for each phecode while covarying for EHR-reported sex, year of birth, median age across the medical records, and the top 10 genetic principal components. We meta-analyzed results of European-like ancestry cohorts from both BioVU and MGBB using fixed-effect models. Phecodes were required to be available at both sites, with a minimum number of cases of 100, yielding 1,520 phecodes. Bonferroni-corrected thresholding (α=0.05/(1520*6)=5.48e-6) was applied to account for the number of phecodes and factors tested. Because phecodes are correlated, we also provide the less stringent false discovery rate (FDR) correction. To examine whether certain disease categories were statistically overrepresented, hypergeometric tests were performed based on the number of significant associations identified per disease category compared to the other categories.

Results were reported as fold enrichment (odds ratio) and were adjusted for the number of categories and factors using FDR correction.

#### LabWAS for genomic factor PGSs

For each site, laboratory test results collected during routine clinical care were extracted from the respective EHR and cleaned following the established QualityLab pipeline^10^. All labs retained in the current study were quantitative, heritable, and had at least 1000 observations in at least 100 individuals. For each individual, the median lab values were calculated across all post-QC observations, and a rank-based inverse normal transformation was applied to ensure normality. In total, 315 and 221 labs were analyzed from individuals of European-like ancestry in BioVU and MGBB, respectively, and 224 from individuals of African-like ancestry in BioVU. Linear regression models were adjusted for sex, year of birth, median age across the medical record, and the top 10 genetic principal components. Using Logical Observation Identifiers Names and Codes (LOINC) and expert review, we harmonized 123 laboratory measures shared across sites and meta-analyzed (more information in **eMethods**). Bonferroni (α=0.05/(123*6)=6.78e-5) and FDR correction were applied to all associations.

Additionally, we provide complete results from each single site to showcase the site-specific labs (**eFigures 8-9**).

A series of conditional PheWAS and LabWAS models was estimated to help distinguish pleiotropy or mediation from reverse causation or confounding, adjusting sequentially for psychiatric diagnostic status, tobacco use disorder, and body mass index. Full model specifications and results are provided in the **eMethods** and **eResults**.

#### Role of the funding source

The funders had no role in study design, data collection, data analysis, data interpretation, writing of the manuscript, or the decision to submit for publication.

## Results

A total of 121,411 participants were included across 2 health care systems: 66,379 BioVU participants with genetically inferred European-like ancestry (mean±SD age, 49.67±22.23 years; 55.6% female), 13,229 BioVU participants with genetically inferred African-like ancestry (mean±SD age, 39.65±21.31 years; 61.0% female), and 41,803 MGBB participants with genetically inferred European-like ancestry (mean±SD age, 52.26±16.61 years; 54.7% female). Detailed characteristics are provided in **eTable 3**.

### PheWAS meta-analysis in European-like ancestry cohorts

To assess clinical correlates of transdiagnostic genetic risk, we conducted PheWAS meta- analyses for each genomic factor PGS across 1,520 phecodes jointly available in BioVU and MGBB (71 mental health and 1,449 physical health conditions across 17 disease categories). At the Bonferroni threshold (α=5.48×10), all factors showed broad associations, ranging from 29 conditions for the compulsive factor to 221 for the hierarchical factor. FDR correction identified an additional 67–296 associations per factor (732 unique phecodes).

Summary results are shown in **eTable 4**, with full results in **eTables 5.1–5.6**.

Across all genomic factors, associations were most strongly concentrated within mental health phenotypes. Mental disorders showed marked statistical overrepresentation relative to all other disease categories (fold enrichment=6.06–46.12; p_FDR_<4.99×10 ; **eTable 6**), with each factor’s primary psychiatric phenotype among its strongest associations (**Figure 1**).

Although significant associations extended across multiple physical health domains (**eTables 5.1 - 5.6**), enrichment was substantially weaker outside psychiatry (**eTable 6**), suggesting that these genomic factors primarily index psychiatric liability while also exhibiting broader pleiotropy across the medical phenome.

**Endocrine and metabolic conditions** showed the strongest and most differentiated physical health associations across factors (**eFigure 3**). The compulsive factor was inversely associated with cardiometabolic phenotypes, including type 2 diabetes (OR=0.92, 95% CI 0.91–0.94; p=2.2×10 ¹), obesity (0.93, 0.91–0.95; p=1.7×10 ¹), and fluid overload (0.92, 0.89–0.95; p=5.1×10). The thought factor showed inverse associations with autoimmune endocrine disorders, including type 1 diabetes (0.90, 0.87–0.93; p=6.3×10 ¹) and Graves’ disease (0.86, 0.81–0.92; p=2.6×10), though these attenuated after excluding MHC variants (**eTable 5.2**). In contrast, neurodevelopmental, internalizing, and substance use disorder (SUD) factors showed positive associations with endocrine/metabolic phenotypes (11, 6, and 6 significant phecodes, respectively; OR=1.04-1.15).

**Figure 3.**
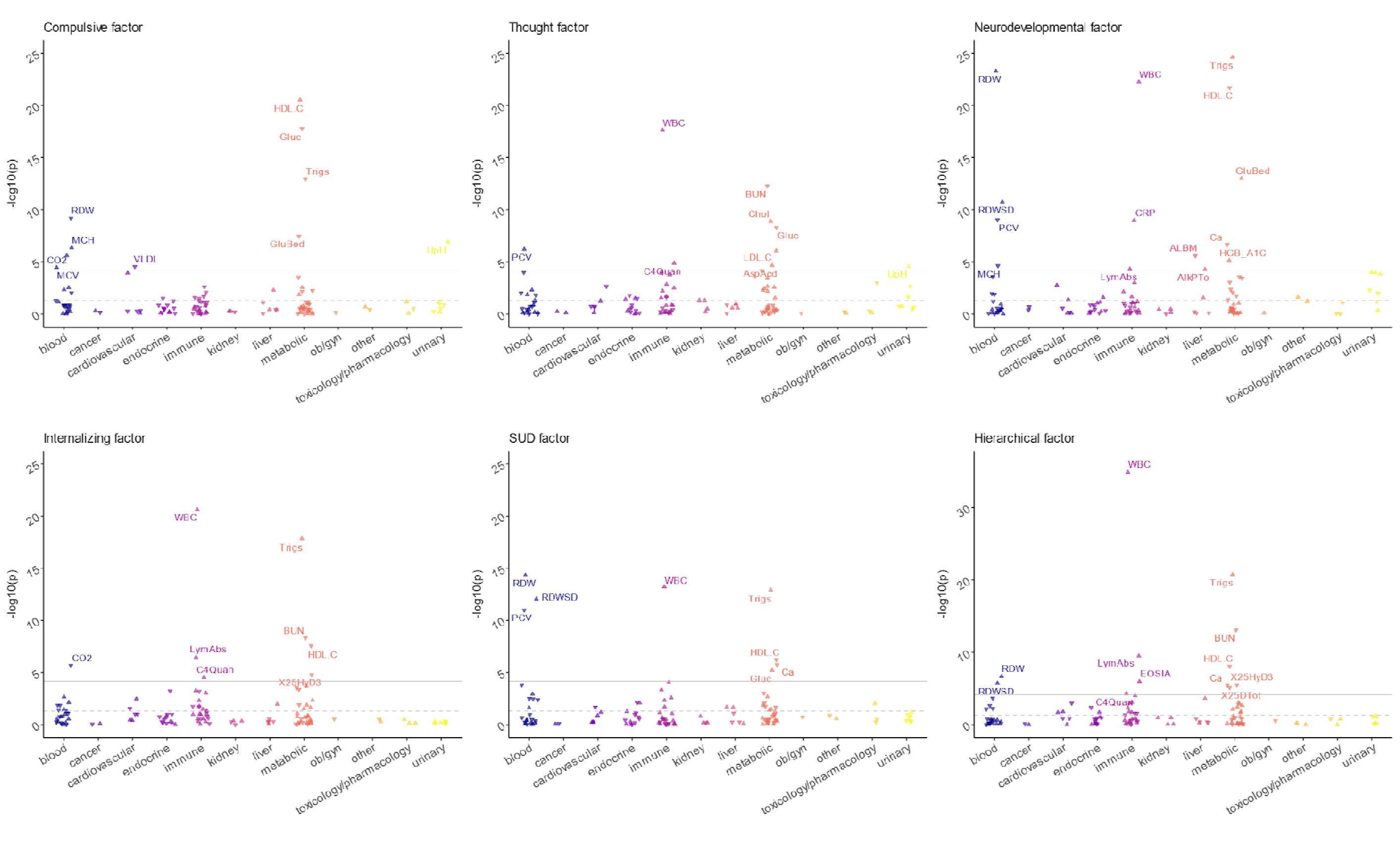
Meta-analyzed laboratory-wide association scan (LabWAS) of PGS for psychiatric genomic factors. A total of 123 labs matched across both si were included. Labs were grouped by system on the x-axis; the significance of each PGS-lab association was shown on the y-axis as -log_10_(p). The direction o triangle indicates the direction of effect. The grey solid line indicates Bonferroni-corrected significance threshold α=0.05/(123*6)=6.78e-5 and the gray dotted indicates α=0.05. Only significant associations are labeled. Full results, including conditional LabWASs, can be found in the interactive eFigures 6**.1-6.6** https://github.com/Yingjie1101/cross-disorder. ALBM, albumin; AlkPTo, alkaline phosphatase; AspAcd, aspartate; BUN, blood urea nitrogen; C4Quan, comp component 4; Ca, calcium; Chol, cholesterol; CO2, carbon dioxide; CRP, C reactive protein; EOSIA, eosinophils; GluBed, bedside glucose; Gluc, glucose; H high-density lipoprotein cholesterol; HGB_A1C, hemoglobin A1c/hemoglobin.total; LDL.C, low-density lipoprotein cholesterol; LymAbs, absolute lymphocy count; MCH, mean corpuscular hemoglobin; MCV, mean corpuscular volume; PCV, hematocrit; RDW, erythrocyte distribution width [Ratio]; RDWSD, erythrocyte distribution width [Entitic volume]; Trigs, triglyceride; UpH, pH of urine; VLDL, very-low-density lipoprotein; WBC, leukocyte (white blood cell) count; X25DTot, 25-Hydroxyvitamin D3+25-Hydroxyvitamin D2; X25HyD3, 25-hydroxyvitamin D3.

**Pain and somatic symptom conditions** were enriched for the thought and internalizing factors (**eTable 6**). The thought factor was associated with abdominal pain (1.04, 1.03–1.06; p=1.6×10), back pain (1.05, 1.03–1.06; p=2.7×10), chest pain (1.04, 1.02–1.05; p=1.8×10), cervicalgia (1.05, 1.03–1.07; p=3.4×10), and fatigue (1.04, 1.03–1.06; p=1.79×10). The internalizing factor showed broader associations, including myalgia (1.13, 1.10–1.16; p=2.7×10 ²), fatigue (1.06, 1.05–1.08; p=2.2×10 ¹), and obstructive sleep apnea (1.08, 1.06–1.10; p=8.6×10 ¹).

**Respiratory and circulatory conditions** were associated with neurodevelopmental, internalizing, and SUD factors. The neurodevelopmental factor was associated with chronic bronchitis (1.16, 1.12–1.21; p=7.7×10 ¹), asthma (1.08, 1.06–1.10; p=9.8×10 ¹), heart failure (1.07, 1.05–1.09; p=1.4×10), hypertension (1.04, 1.03–1.06; p=1.8×10), and myocardial infarction (1.10, 1.07–1.12; p=3.8×10 ¹³). The internalizing factor was associated with asthma (1.08, 1.06–1.10; p=1.6×10 ¹³), chronic airway obstruction (1.09, 1.07–1.12; p=8.8×10 ¹), and shortness of breath (1.07, 1.05–1.08; p=4.0×10 ¹). The SUD factor was associated with chronic bronchitis (1.15, 1.10–1.19; p=1.7×10 ¹²), hypertension (1.06, 1.04–1.07; p=2.8×10 ¹²), and coronary atherosclerosis (1.06, 1.04–1.08; p=1.3×10 ¹).

**Infectious and immune-related conditions** were primarily associated with the thought, neurodevelopmental, and SUD factors. Shared associations included viral hepatitis (OR=1.13–1.26) and HIV (1.17–1.23). The thought factor was additionally associated with urinary tract infection (1.04, 1.03–1.06; p=1.6×10), and the SUD factor with pneumonia (1.07, 1.05–1.09; p=4.7×10 ¹¹).

Additional associations included positive links between the internalizing factor and gastrointestinal disorders, including gastroesophageal reflux disease (1.09, 1.07–1.10; p=2.6×10 ³) and esophageal disease (1.09, 1.07–1.10; p=1.9×10 ²), as well as inverse associations between the neurodevelopmental factor and dermatologic phenotypes, including benign skin neoplasms (0.91, 0.90–0.93; p=3.5×10 ²²) and dyschromia (0.92, 0.90–0.94; p=7.0×10 ¹³).

Conditional PheWAS showed that most associations were robust to adjustment for psychiatric diagnoses, tobacco use disorder, and BMI (**eTables 7.1–7.6**; **eFigures 2.1–2.6**).

### PheWAS in the BioVU African-like ancestry cohort

We additionally conducted PheWAS in the BioVU African-like ancestry cohort (n=13,203) across 724 phecodes with ≥100 cases. Although no associations reached Bonferroni significance (**eFigure 5**), 81.4–97.1% of effect directions were concordant with the European-like ancestry meta-analysis, indicating broadly similar patterns despite reduced power. Several associations reached FDR significance, including a negative association between the compulsive factor and tobacco use disorder (OR=0.89, 0.84–0.94; p=5.3×10), not observed in European-like ancestry, a positive association of the SUD factor with hypotension (1.17, 1.08–1.27; p=0.001) and tobacco use disorder (1.15, 1.08– 1.22; p=1.3×10), and a positive association of the hierarchical factor with mood disorders (1.12, 1.06–1.19; p=1.5×10).

To enhance readability, only top associations are labeled. Full results (including conditional PheWASs) can be found in the interactive **eFigures 2.1-2.6** https://github.com/Yingjie1101/cross-disorder. SUD: substance use disorders.

### LabWAS meta-analysis in European-like ancestry cohorts

LabWAS meta-analyses across 123 laboratory measures identified broad physiological correlates of the genomic factors (**eTable 8**). Across factors, 27 measures surpassed the Bonferroni threshold (α=6.78×10), and 61 met FDR criteria (**Figure 3**, **eTables 9.1–9.6**). Most associations involved metabolic, hematologic, or immune-related markers, consistent with patterns observed in the PheWAS analyses.

**Metabolic measures** showed some of the strongest and most differentiated associations across factors. Triglyceride levels were inversely associated with the compulsive factor (β[SE]=−0.034[0.005], p=1.2×10 ¹³) but positively associated with the neurodevelopmental, internalizing, SUD, and hierarchical factors (β=0.034–0.047). HDL cholesterol showed the opposite pattern, with positive associations for the compulsive factor (β[SE]=0.041[0.004], p=3.1×10 ²¹) and inverse associations for the other first-order factors (β=−0.042 to −0.022), with no significant association for the thought factor. Blood glucose also showed divergent associations, being positively associated with the neurodevelopmental and SUD factors (β[SE]=0.034[0.003], p=7.1×10 ² ; β[SE]=0.015[0.003], p=6.2×10) and inversely associated with the compulsive and thought factors (β[SE]=−0.028[0.003], p=1.7×10 ¹ ; β[SE]=−0.019[0.003], p=5.1×10).

Immune and hematologic markers showed broad cross-factor associations. White blood cell count (WBC) was positively associated with the thought, neurodevelopmental, internalizing, SUD, and hierarchical factors (β=0.023–0.038), and inversely associated with the compulsive factor at FDR significance (β=−0.010).

Complement C4 was associated with the thought, and internalizing factors (β=0.052–0.055), while C-reactive protein (CRP) was associated with the neurodevelopmental factor (β[SE]=0.038[0.01], p=1.1×10). Hematocrit showed inverse associations with neurodevelopmental and SUD factors (β=−0.019 and −0.022) but a positive association with the thought factor (β[SE]=0.016[0.003], p=6.2×10). Erythrocyte distribution width (RDW) showed the opposite pattern, with positive associations for neurodevelopmental and SUD factors (β=0.033 and 0.026) and an inverse association for the compulsive factor (β[SE]=−0.020[0.003], p=6.6×10 ¹).

Additional site-specific laboratory measures unavailable for meta-analysis, including creatinine, nucleated erythrocytes, and calcitriol, showed several significant associations (**Figure 4**; **eFigures 8–9**), with some remaining after full model adjustment.

**Figure 4.**
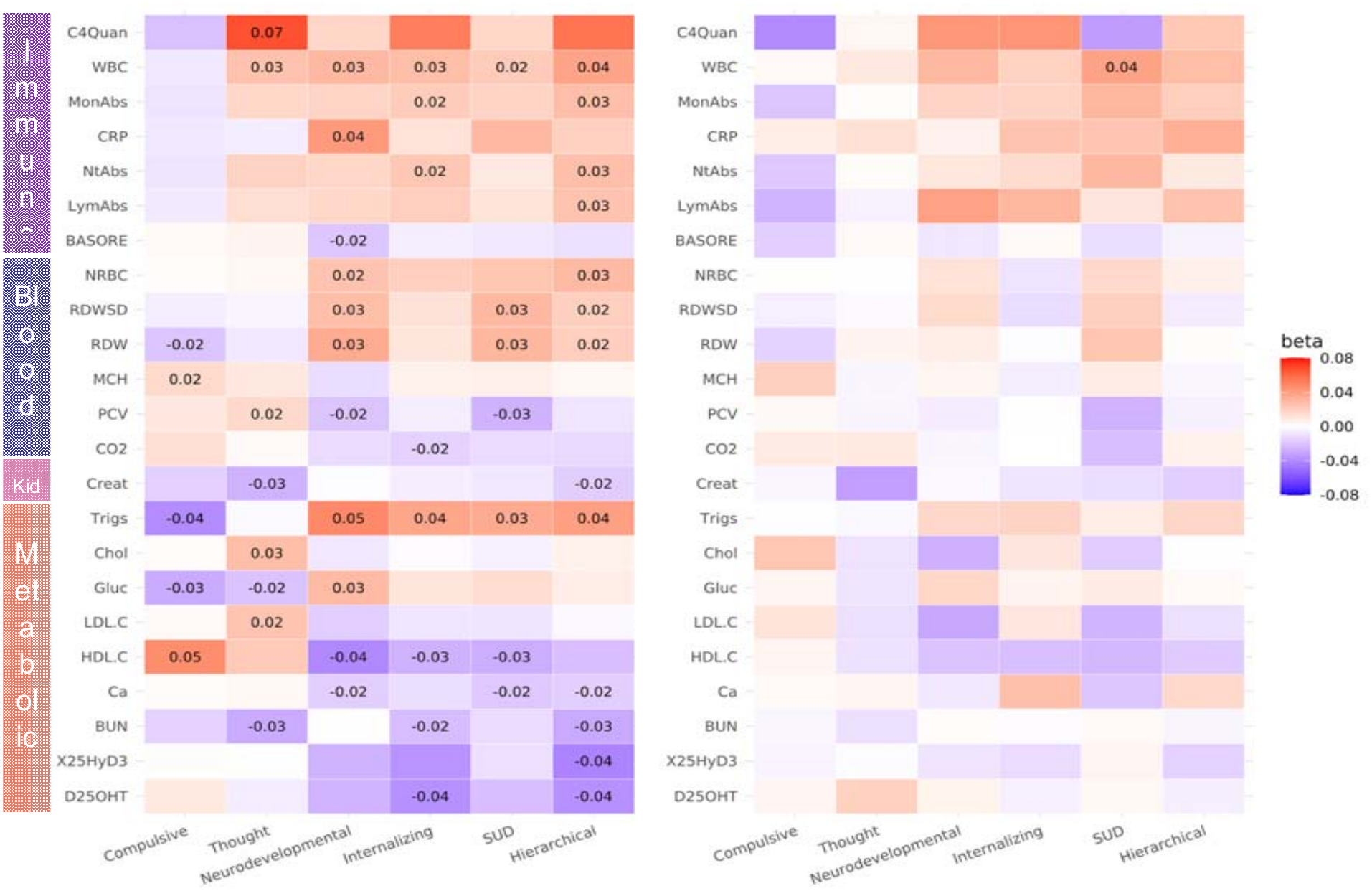
PGS-lab associations for BioVU samples of European-like ancestry (left) and African-like ancestry (right). Standardized effect sizes (β) of significant PGS-lab associations are annotated. C4Quan, complement component 4; WBC, leukocyte (white blood cell) count; MonAbs, absolute monocyte count; CRP, C reactive protein; NtAbs, absolute neutrophil count; LymAbs, absolute lymphocytes count; BASORE, basophils/100 leukocytes ratio; NRBC, nucleated erythrocytes/100 leukocytes ratio; RDWSD, erythrocyte distribution width [Entitic volume]; RDW, erythrocyte distribution width [Ratio]; MCH, mean corpuscular hemoglobin; PCV, hematocrit; CO2, carbon dioxide; Creat, creatinine; Trigs, triglyceride; Chol, cholesterol; Gluc, glucose; LDL.C, low-density lipoprotein cholesterol; HDL.C, high-density lipoprotein cholesterol; Ca, calcium; BUN, blood urea nitrogen; X25HyD3, 25-hydroxyvitamin D3; D25OHT, Vitamin D+Metabolites.

### LabWAS in the BioVU African-like ancestry cohort

In the BioVU African-like ancestry cohort (**Figure 4**), fewer measures reached significance, likely reflecting reduced statistical power. WBC was positively associated with the SUD factor at the Bonferroni threshold (β[SE]=0.038[0.009], p=3.1×10), and creatinine was inversely associated with the thought factor at FDR significance (β[SE]=−0.033[0.009], p=0.002). Despite limited power, 74.6% of associations showed concordant effect directions with the European-like ancestry analyses, with the highest concordance for the neurodevelopmental factor (87.0%) and lowest for the thought factor (56.5%).

## Discussion

To our knowledge, this is the first study to comprehensively investigate associations between genetically defined transdiagnostic psychiatric factors and a broad spectrum of medical diagnoses and laboratory biomarkers across large electronic health record-linked biobanks. Leveraging data from more than 120,000 individuals from two U.S. health care systems, we examined liabilities for six transdiagnostic psychiatric factors using both PheWAS and LabWAS approaches. At the level of diagnoses (PheWAS), results showed partial specificity within the psychiatric domain, with stronger associations for primary psychiatric phenotypes relative to other mental disorders, alongside widespread associations across physical health conditions. Different physiological systems were implicated by different factors, particularly endocrine/metabolic, circulatory, respiratory, and digestive systems. At the level of lab-based markers (LabWAS), results reinforced these patterns by identifying convergent associations with lipid and glucose metabolism, hematologic traits, and immune and inflammatory markers.

In keeping with the shared genetic architecture across psychiatric dimensions, we identified overlapping physiological systems across factors. The internalizing, neurodevelopmental, and SUD factors, three of the most correlated factors, also shared the greatest phenotypic overlap, with associations spanning circulatory, respiratory, and digestive systems.

We detected divergent associations with metabolic disease across the genetically defined psychiatric factors: internalizing, neurodevelopmental, and SUD factors were associated with increased risks for type 2 diabetes, obesity, and poor lipid metabolism profiles even after adjustment for psychiatric diagnoses, showing consistent results with previous reports^12^. Secondly, the compulsive factor was associated with decreased risks for obesity, type 1 and type 2 diabetes, and with enhanced glucose and lipid metabolism. Similarly, the thought factor was associated with decreased risk for type 1 diabetes and type 2 diabetes (only with FDR correction), and with increased levels of LDL cholesterol and decreased levels of glucose. Such ‘protective’ genetic effects may seem to be contradictory to epidemiological studies reporting increased prevalence of metabolic diseases in OCD^13^ and SCZ^14^, where the effects of genetic predisposition could be obscured by other disorder-related non-genetic influences such as side effects of antidepressant^15^ or antipsychotic medications^16^ and/or correlated environmental factors and lifestyle factors. This is further supported by the observation that the negative associations became stronger after controlling for diagnostic status (**eFigure 4**, **eTable 7.1-7.2**). Similarly, the negative association between genetic risks for neurodevelopmental disorders and dermatologic conditions, observed here and in prior PheWAS studies,^17,18^ may contrast with epidemiological comorbidities (e.g., ADHD and skin disorders^19,20^), potentially reflecting behavioral rather than genetic mechanisms.

Several laboratory correlates of the psychiatric factors linked to altered immune and inflammatory functions, align with emerging immuno-psychiatry research. Building on prior PsycheMERGE findings^10^ linking depression-PGS with WBC, we observed associations with inflammatory markers across all factors except the compulsive factor. These associations feature both innate immunity markers such as monocytes and neutrophils, as well as lymphocytes, which are central to adaptive immunity, mirroring observations from case-control cohort studies.^21,22^ Previous studies have reported inconsistent findings regarding the relationship between psychiatric disorders and CRP,^23^ an acute-phase protein crucial for diagnosing and monitoring various medical conditions. The association with CRP was captured strongest through the neurodevelopmental factor, accompanied by decreased albumin levels, supporting recent findings linking the CRP-to-albumin ratio to an increased risk for psychiatric disorders.^21^ Chronic low-grade inflammation can sustain systemic immune activation, contributing to elevated risks for conditions like diabetes, autoimmune diseases, and atherosclerosis. This chronic inflammation, marked by elevated CRP and proinflammatory cytokines from neutrophils and monocytes, may lead to neuroinflammation by crossing the blood-brain barrier (BBB). This process activates microglia and astrocytes, which are increasingly considered to play pathogenic roles in psychiatric disorders.^24^ We also demonstrated an association between the thought factor and elevated complement C4 levels in peripheral blood, aligning with research linking higher levels of C4A expression to excessive synaptic pruning in the brain, a process implicated in schizophrenia pathogenesis.^25^ We additionally observed negative associations between the thought factor and autoimmune diseases such as Type 1 diabetes, Celiac, and Graves’ disease, all of which were also linked to the MHC region.

Given the complex genetic architecture of this region and inconsistent previous findings ^26–28^ further investigations of the genetic and immunological interactions underlying the thought factor are warranted.

This study is the first, to the best of our knowledge, that comprehensively assessed genetically defined transdiagnostic psychiatric factors across medical diagnoses and laboratory markers derived from clinical settings. Transdiagnostic PGS may offer greater power to identify shared genetic associations across disorders, whereas disorder-specific PGS may be more informative for identifying associations unique to individual diagnoses. We identified up to 732 unique medical conditions and 61 lab measures linked to psychiatric risks, with PheWAS and LabWAS providing converging evidence for involvement of specific physiological systems. The association patterns were broadly consistent with results from previous single-disorder PGS-PheWAS (e.g., for SCZ^29^, MDD^17,30^, PTSD^31^) and -LabWAS (e.g., for SCZ, BP, and MDD^32^, and PTSD^31^) studies, supporting the validity of the multivariate factor structure. In addition to single-disorder-based PGSs, one previous PheWAS study^18^ incorporated a cross-disorder factor based on five psychiatric disorders (ASD, ADHD, MDD, BP, and SCZ) and identified a single significant association with mood disorders. Our results confirmed this association, and, importantly, uncovered hundreds of additional links between mental and physical conditions. Furthermore, a recent study in a different biobank sample employed a complementary approach through shared factor modeling for psychiatric and somatic conditions; they also observed widespread sharing of risk factors between mental and somatic domains. Compared to this study, our scope of discovery was enhanced by using cross-site sample curation and harmonization of two large biobanks, a cross-ancestry approach, and the combination of PheWAS and LabWAS, highlighting clinically relatable brain-body associations at the diagnostic and laboratory marker level.

Besides the strengths and novelty of this work, several limitations should also be noted. First, the GWAS discovery samples lacked ancestral diversity, resulting in genomic factor structures derived from individuals of European-like genetic ancestry. This limitation reduced the predictive power in other ancestral groups. Although the African-like ancestry sample in BioVU showed broadly similar association patterns, larger and more diverse cohorts are needed to improve generalizability. Second, despite significant efforts to harmonize protocols and standardize phenotypic definitions, differences in demographic characteristics and disease prevalence between the sites contributed to heterogeneity. For example, 44%-61% of laboratory measures were not shared between the sites, even after rank-based matching. To address this, we provide between-site heterogeneity statistics for all phenotypes and report single-site LabWAS results to highlight site-specific lab tests. Thirdly, the LabWAS results may be influenced by selection bias and bias by indication, as laboratory testing in EHR settings is driven by clinical need rather than systematic population sampling. Additionally, although many associations reached statistical significance, their effect sizes were generally small, suggesting limited clinical relevance at the individual level. Nevertheless, these associations may provide insights into the shared biological mechanisms underlying psychiatric and somatic conditions. Finally, while our genetic approach provides a means to explore transdiagnostic psychiatric dimensions, caution is needed when interpreting these associations, given the complex interplay between genetic, environmental, and behavioral factors. Additionally, because GWAS summary statistics continue to be updated as larger datasets become available, future studies may refine the genomic factor structure and uncover additional associations. Nevertheless, the concordance of our findings with recent studies using more up-to-date GWAS data suggests that the principal dimensions and relationships identified here are likely to be robust, with larger datasets primarily increasing power to detect additional signals. Our findings underscore the need for further research to clarify the interplay between genetic predispositions and the phenotypic manifestations of these dimensions.

## Conclusions

Using genetic modelling across the psychopathology spectrum, this study underscored the close link between mental and somatic health. The associations span multiple organ systems, with notable involvement of metabolic and immune pathways. These findings support ongoing efforts to define biologically informed subtypes of psychiatric disorders, such as immuno-metabolic profiles, which can facilitate patient stratification in clinical trials and support transdiagnostic precision psychiatry approaches. At the same time, the level of evidence now available for immune/metabolic involvement in mental health- coming from complementary study designs - should raise clinicians’ awareness to monitor physical health in psychiatry and vice versa. Our findings clearly show that such a need for monitoring goes beyond individual psychiatric conditions, supporting the development of transdiagnostic protocols and guidelines.

## Funding/acknowledgements

The work was supported by the funding from the institutes involved. BF’s contribution was supported by funding from the European Community’s Horizon 2020 Programme (H2020/2014 – 2020) under grant agreement n° 847879 (PRIME). Research reported in this publication was also supported by the National Institute Of Mental Health of the National Institutes of Health under Award Number R01MH124851. The content is solely the responsibility of the authors and does not necessarily represent the official views of any of the funders. LKD was supported by a grant from the National Institutes of Health R01MH118223. ES is funded by a Horizon Europe Research and Innovation Program (FAMILY; Grant agreement 101057529) and ERC Consolidator Grant (EM- POWER, project number 101170512), funded by the European Union. Views and opinions expressed are however those of the author(s) only and do not necessarily reflect those of the European Union or the European Research Council. Neither the European Union nor the granting authority can be held responsible for them.

## Synthetic Derivative

The project described was supported by the National Center for Research Resources, Grant UL1 RR024975-01, and is now at the National Center for Advancing Translational Sciences, Grant 2 UL1 TR000445-06. The content is solely the responsibility of the authors and does not necessarily represent the official views of the NIH.

## BioVU

The dataset(s) used for the analyses described were obtained from Vanderbilt University Medical Center’s BioVU which is supported by numerous sources: institutional funding, private agencies, and federal grants. These include the NIH funded Shared Instrumentation Grant S10RR025141; and CTSA grants UL1TR002243, UL1TR000445, and UL1RR024975. Genomic data are also supported by investigator-led projects that include U01HG004798, R01NS032830, RC2GM092618, P50GM115305, U01HG006378, U19HL065962, R01HD074711; and additional funding sources listed at https://victr.vumc.org/biovu-funding/.

## Data sharing

EHR data cannot be made publicly available due to institutional and ethical restrictions governing the participating biobanks. Analysis scripts and summary-level results will be made available through a public GitHub repository upon publication.

## Author contributions

YS, ES, NRM, BF, and LKD conceptualized and designed the study. YS, YZ, YHL, PS, FB, TG performed the statistical analyses and verified the underlying data. YS, YHL, PS, FB, YZ, contributed to data curation, and interpretation. YS drafted the manuscript, with contributions from MN, LKD, BF, ES, NRM. All authors critically revised the manuscript, approved the final version, and accept responsibility for the decision to submit for publication.

## Declaration of interests

BF has received educational speaking fees and travel support from Medice GmbH. All other authors report no potential conflicts of interest.

## Declaration of generative AI and AI-assisted technologies in the manuscript preparation process

During the preparation of this work, YS and MN used ChatGPT for grammar and spelling checks as well as reducing word count. The authors reviewed and edited the output as needed and take full responsibility for the content of the manuscript.

## Supporting information

eTables

eMethods

## Data Availability

https://github.com/Yingjie1101/cross-disorder

## Notes

### Author Declarations

The VUMC Institutional Review Board approved this project (IRB#172020). The participants in the Mass General Brigham Biobank (MGBB) were recruited during an in-person interaction with clinical research staff, through an electronic consent workflow, or through collaborating studies. The study was approved by the Institutional Review Board (or Ethics Committee) of Mass General Brigham (protocol code 2009-P-002312). Informed consent was obtained from all patients. In both biobanks, the genomic sample of each participant can be linked with extensive electronic health record data and clinical laboratory test results.

## References

1. Firth, J. et al. The Lancet Psychiatry Commission: a blueprint for protecting physical health in people with mental illness. The Lancet Psychiatry 6, 675–712 (2019).

2. Simmons, J. M. & Quinn, K. J. The NIMH Research Domain Criteria (RDoC) Project: implications for genetics research. Mamm Genome 25, 23–31 (2014).

3. Kotov, R. et al. The hierarchical Taxonomy of Psychopathology (HiTOP): A dimensional alternative to traditional nosologies. J Abnorm Psychol 126, 454–477 (2017).

4. Grotzinger, A. D. et al. Transcriptome-Wide Structural Equation Modeling of 13 Major Psychiatric Disorders for Cross-Disorder Risk and Drug Repurposing. JAMA Psychiatry (2023) doi:10.1001/jamapsychiatry.2023.1808.

5. Murray GK, Lin T, Austin J, McGrath JJ, Hickie IB, Wray NR. Could Polygenic Risk Scores Be Useful in Psychiatry?: A Review. JAMA Psychiatry. 2021 Feb 1;78(2):210–219. doi: 10.1001/jamapsychiatry.2020.3042.

6. Denny, J. C., Bastarache, L. & Roden, D. M. Phenome-Wide Association Studies as a Tool to Advance Precision Medicine. Annu Rev Genomics Hum Genet 17, 353–373 (2016).

7. Bush, W. S., Oetjens, M. T. & Crawford, D. C. Unravelling the human genome–phenome relationship using phenome-wide association studies. Nat Rev Genet 17, 129–145 (2016).

8. Dennis, J. K. et al. Clinical laboratory test-wide association scan of polygenic scores identifies biomarkers of complex disease. Genome Med 13, 6 (2021).

9. Ge, T., Chen, C.-Y., Ni, Y., Feng, Y.-C. A. & Smoller, J. W. Polygenic prediction via Bayesian regression and continuous shrinkage priors. Nat Commun 10, 1776 (2019).

10. Sealock, J. M. et al. Use of the PsycheMERGE Network to Investigate the Association Between Depression Polygenic Scores and White Blood Cell Count. JAMA Psychiatry 78, 1365–1374 (2021).

11. Denny, J. C. et al. PheWAS: demonstrating the feasibility of a phenome-wide scan to discover gene- disease associations. Bioinformatics 26, 1205–1210 (2010).

12. Fanelli, G. et al. Insulinopathies of the brain? Genetic overlap between somatic insulin-related and neuropsychiatric disorders. Transl Psychiatry 12, 1–8 (2022).

13. Isomura, K. et al. Metabolic and Cardiovascular Complications in Obsessive-compulsive Disorder: A Total Population, Sibling Comparison Study With Long-Term Follow-up. Biological Psychiatry 84, 324–331 (2018).

14. Mitchell, A. J. et al. Prevalence of Metabolic Syndrome and Metabolic Abnormalities in Schizophrenia and Related Disorders—A Systematic Review and Meta-Analysis. Schizophrenia Bulletin 39, 306–318 (2013).

15. Lee, S. H., Paz-Filho, G., Mastronardi, C., Licinio, J. & Wong, M.-L. Is increased antidepressant exposure a contributory factor to the obesity pandemic? Transl Psychiatry 6, e759–e759 (2016).

16. De Hert, M., Detraux, J., van Winkel, R., Yu, W. & Correll, C. U. Metabolic and cardiovascular adverse effects associated with antipsychotic drugs. Nat Rev Endocrinol 8, 114–126 (2012).

17. Fang, Y., Fritsche, L. G., Mukherjee, B., Sen, S. & Richmond-Rakerd, L. S. Polygenic Liability to Depression Is Associated With Multiple Medical Conditions in the Electronic Health Record: Phenome-wide Association Study of 46,782 Individuals. Biological Psychiatry 92, 923–931 (2022).

18. Kember, R. L. et al. Polygenic Risk of Psychiatric Disorders Exhibits Cross-trait Associations in Electronic Health Record Data From European Ancestry Individuals. Biol Psychiatry 89, 236–245 (2021).

19. Instanes, J. T., Klungsøyr, K., Halmøy, A., Fasmer, O. B. & Haavik, J. Adult ADHD and Comorbid Somatic Disease: A Systematic Literature Review. J Atten Disord 22, 203–228 (2018).

20. Akmatov, M. K., Ermakova, T. & Bätzing, J. Psychiatric and Nonpsychiatric Comorbidities Among Children With ADHD: An Exploratory Analysis of Nationwide Claims Data in Germany. J Atten Disord 25, 874– 884 (2021).

21. Zeng, Y. et al. Inflammatory Biomarkers and Risk of Psychiatric Disorders. JAMA Psychiatry (2024) doi:10.1001/jamapsychiatry.2024.2185.

22. Milaneschi, Y. et al. Association of inflammation with depression and anxiety: evidence for symptom- specificity and potential causality from UK Biobank and NESDA cohorts. Mol Psychiatry 26, 7393–7402 (2021).

23. Misiak, B. et al. Peripheral blood inflammatory markers in patients with attention deficit/hyperactivity disorder (ADHD): A systematic review and meta-analysis. Progress in Neuro-Psychopharmacology and Biological Psychiatry 118, 110581 (2022).

24. Najjar, S., Pearlman, D. M., Alper, K., Najjar, A. & Devinsky, O. Neuroinflammation and psychiatric illness. Journal of Neuroinflammation 10, 816 (2013).

25. Sekar, A. et al. Schizophrenia risk from complex variation of complement component 4. Nature 530, 177– 183 (2016).

26. Chen, M.-H. et al. Type 1 diabetes mellitus and risks of major psychiatric disorders: A nationwide population-based cohort study. Diabetes & Metabolism 48, 101319 (2022).

27. Hallford, P., Clair, D. S., Halley, L., Mustard, C. & Wei, J. A study of type-1 diabetes associated autoantibodies in schizophrenia. Schizophrenia Research 176, 186–190 (2016).

28. Juvonen, H. et al. Incidence of Schizophrenia in a Nationwide Cohort of Patients With Type 1 Diabetes Mellitus. Archives of General Psychiatry 64, 894–899 (2007).

29. Zheutlin, A. B. et al. Penetrance and Pleiotropy of Polygenic Risk Scores for Schizophrenia in 106,160 Patients Across Four Health Care Systems. Am J Psychiatry 176, 846–855 (2019).

30. Mulugeta, A., Zhou, A., King, C. & Hyppönen, E. Association between major depressive disorder and multiple disease outcomes: a phenome-wide Mendelian randomisation study in the UK Biobank. Mol Psychiatry 25, 1469–1476 (2020).

31. Pathak, G. A. et al. Genetic Liability to Posttraumatic Stress Disorder Symptoms and Its Association With Cardiometabolic and Respiratory Outcomes. JAMA Psychiatry 81, 34–44 (2024).

32. Sewell, M. D. E. et al. Associations between major psychiatric disorder polygenic risk scores and blood- based markers in UK biobank. *Brain*, Behavior, and Immunity 97, 32–41 (2021).

