## Supplementary material for "Transdiagnostic Psychiatric Polygenic Scores Reveal Pleiotropic Effects Across Medical and Laboratory Phenome": eMethods

**Supplementary Information**

### **eMethods**

**eResults**

### **eFigure 1. Multivariate genetic structures of 13 psychiatric disorders.**

### **eFigure 2. Meta-analyzed primary and step-wise conditional PheWAS results.**

### [eFigure 2.1: Compulsive-PGS PheWAS](https://yingjie1101.github.io/cross-disorder/PheWAS_Comp.html)

### [eFigure 2.2: Thought-PGS PheWAS](https://yingjie1101.github.io/cross-disorder/PheWAS_Thou.html)

### [eFigure 2.3: Neurodevelopmental-PGS PheWAS](https://yingjie1101.github.io/cross-disorder/PheWAS_Neur.html)

### [eFigure 2.4: Internalizing-PGS PheWAS](https://yingjie1101.github.io/cross-disorder/PheWAS_Inte.html)

### [eFigure 2.5: SUD-PGS PheWAS](https://yingjie1101.github.io/cross-disorder/PheWAS_SUD.html)

### [eFigure 2.6: Hierarchical-PGS PheWAS](https://yingjie1101.github.io/cross-disorder/PheWAS_Hier.html)

### **eFigure 3. The distribution of odds ratios (OR) for somatic disease categories across genomic factor PGSs after full model adjustment.**

### **eFigure 4. Top instances of associations with reversed effect size change in conditional PheWAS.**

### **eFigure 5. PheWAS of PGS for psychiatric genomic factors in BioVU African cohort.**

### **eFigure 6. Meta-analyzed primary and step-wise conditional LabWAS results.**

### [eFigure 6.1: Compulsive-PGS LabWAS](https://yingjie1101.github.io/cross-disorder/LabWAS_Comp.html)

### [eFigure 6.2: Thought-PGS LabWAS](https://yingjie1101.github.io/cross-disorder/LabWAS_Thou.html)

### [eFigure 6.3: Neurodevelopmental-PGS LabWAS](https://yingjie1101.github.io/cross-disorder/LabWAS_Neur.html)

### [eFigure 6.4: Internalizing-PGS LabWAS](https://yingjie1101.github.io/cross-disorder/LabWAS_Inte.html)

### [eFigure 6.5: SUD-PGS LabWAS](https://yingjie1101.github.io/cross-disorder/LabWAS_SUD.html)

### [eFigure 6.6: Hierarchical-PGS LabWAS](https://yingjie1101.github.io/cross-disorder/LabWAS_Hier.html)

**eFigure 7. Effect sizes in primary and conditional LabWAS models.**

### **eFigure 8. Laboratory-wide association scan (LabWAS) of PGS for psychiatric genomic factors at BioVU.**

### **eFigure 9. Laboratory-wide association scan (LabWAS) of PGS for psychiatric genomic factors at MGBB.**

### **eReferences**

### **eMethods**

#### **Genotyping and quality control**

Vanderbilt University Medical Center biobank (BioVU)

Details of the genotyping and quality control processes for BioVU samples have been described previously.^1,2^ Briefly, genotyped data were obtained using the Illumina MultiEthnic Genotyping Array (MEGAEX) and filtered for sample and SNP call rates, sex mismatch, and heterozygosity. Imputation was carried out using the Michigan Imputation Server with the Haplotype Reference Consortium reference panel. Individuals of European or African ancestry were identified using a series of principal component analyses (PCA) implemented in Eigenstrat, following previously established procedures.^2^ After post-imputation quality control, the final dataset contained 6,360,980 SNPs on 66,917 individuals of genetically defined European-like ancestry and 28,746,646 SNPs on 13,329 individuals of of genetically defined African-like ancestry.

Mass General Brigham Biobank (MGBB)

Details of the genotyping and quality control processes for MGBB samples have been described previously on Github (GSA samples: <https://github.com/getian107/MGBB-QC>, MEGA samples: <https://github.com/Annefeng/PBK-QC-pipeline>). Briefly, the first subset of MGBB sample (GSA samples) were genotyped on Global Screening Array (GSA) kits from Illumina and the second subset of MGBB sample (MEGA sample) were genotyped using the Multi-Ethnic Global Array (MEGA) kits from Illumina. These genotyped data were filtered for sample and SNP call rates, sex mismatch, and heterozygosity. Imputation was carried out using the Michigan Imputation Server with the Haplotype Reference Consortium reference panel. Individuals of European ancestry were identified using a series of principal component analyses (PCA) as described on Github. After post-imputation quality control, the final dataset of European genetic ancestry contained 8,679,265 SNPs on 33,067 individuals from the GSA sample and 7,353,845 SNPs on 25,698 individuals from the MEGA sample. The GSA sample and MEGA sample have a sample overlap of 15,442 individuals. In our analyses we deduplicated the sample overlap by prioritizing the GSA sample because of the higher number of overlapping SNPs with those contributing to PRS. In the analytical sample, we included 31,948 GSA samples and 9,855 MEGA samples with lab records in their electronic health records.

#### **Psychiatric genomic factors**

We adopted both the correlated factors model and the hierarchical factor model based on results from Grotzinger et al. (2023)^3^. Briefly, thirteen univariate psychiatric disorders were included to construct the genomic structure equation models, namely anorexia nervosa (AN), anxiety disorders (ANX), attention-deficit/hyperactivity disorder (ADHD), autism spectrum disorder (AUT), bipolar disorder (BIP), cannabis use disorder (CUD), major depressive disorder (MDD), obsessive-compulsive disorder (OCD), opioid use disorder (OUD), post-traumatic stress disorder (PTSD), alcohol use disorder (AUD), schizophrenia (SCZ), and Tourette syndrome (TS). The total sample sizes for each univariate GWAS range from 9,537 to 500,199. Details of all contributing cohorts and the corresponding GWAS studies were provided in **eTable 1**. The genetic covariance matrix was estimated using the *LDSC* function within the *GenomicSEM* package in R and genetic correlations among 13 psychiatric disorders were presented in **eFigure 1**.

The correlated factors model consists of five factors that are inter-correlated with each other, whereas the hierarchical factor model further models the inter-factor correlations with a higher-order general psychopathology factor (i.e., the Hierarchical factor). Both of the models fit the data well (correlated factors model: CFI=0.95, SRMSR=0.102; hierarchical factor model CFI=0.900; SRMSR=0.129), and were carried forward for multivariate GWASs.

Polygenic scores were constructed using PRS-CS^4^, which utilizes a Bayesian-based framework to infer single nucleotide polymorphism (SNP) effect sizes by applying continuous shrinkage. The European and African references from the 1000 Genomes Project phase 3 were used as the LD reference panels, respectively. Given the highly complex linkage disequilibrium (LD) structure within the major histocompatibility complex region (MHC), we performed a set of sensitivity analyses by recalculating PGSs after removing 9,446 SNPs within this region (chr6:25Mb-34Mb) and estimated the change in associations compared to the primary set of PGSs.

As a sensitivity analysis, we excluded the SNPs that had divergent effects across the indicators (i.e., disorders in the correlated factors model and factors in the hierarchical model) before the polygenic score analysis. The SNPs were identified based on the Q_SNP_ heterogeneity metric, which is the χ^2^ difference between a model in which the SNP operates through the latent factor and a model in which the SNP operates directly on the indicators. The Q_SNP_ metric of the correlated factors model indicates the SNPs with disorder-specific effects, and the Q_SNP_ metric of the hierarchical model indicates the SNPs with factor-specific effects. We removed the SNPs with significant Q_SNP_ effects from the summary statistics of each factor, along with the SNPs in linkage disequilibrium (r^2^ > 0.1, 500 kb window^5^) with them. This has resulted in minimal change in the results, therefore we only reported the results with PGS constructed based on the complete sets of SNPs in the main text.

#### **Mapping and harmonization of laboratory tests across BioVU and MGBB sites**

We provided an overview of the mapping of labs across sites in **eTable 8**. Complete sets of laboratory tests from BioVU and MGBB were initially matched based on exact LOINC codes, ensuring that identical tests were paired automatically (exact_match=TRUE). For tests without a direct LOINC match, a manual matching process was undertaken to verify and ensure the correspondence between lab tests across the two sites. Given that a rank-based inverse normal transformation (INT) was applied to all laboratory values as part of the QualityLab pipeline, we ensured that at least a rank-based correspondence was maintained between the paired labs. This approach was implemented to maximize the overlap and comparability of test results across both cohorts and was cross-verified by a clinical chemist with expertise in laboratory medicine. For consistency, we reference the labs with the naming and system assignment at the BioVU site, but full details of the lab can be retrieved from **eTable 8**. Additionally, we included site-specific results in **eFigure 8** and **eFigure 9** to showcase non-overlapping labs.

#### **Sensitivity analyses**

A series of conditional PheWAS and LabWAS models was estimated to help distinguish pleiotropy and/or mediation from reverse causation and/or confounding. In addition to baseline covariates adjusted in the primary models, we further derived case-control coding for psychiatric diagnostic status, both factor-specifically and factor-generally. The factor-specific diagnoses encoded the index diagnoses that were either the primary or closely-related phenotypes to the factor in the multivariate GWAS (**eTable 2**). Factor-specific diagnoses captured the primary or closely related disorders contributing to each genomic factor, whereas factor-general diagnoses reflected any psychiatric disorder represented across factors. We additionally adjusted sequentially for tobacco use disorder (phecode 307), which is not included in any factor, and for body mass index (BMI).

### **eResults**

#### **PheWAS: secondary trait–adjusted models**

To assess the impact of secondary traits linked with the psychiatric genetic risk on the PGS-phenotype associations, we ran PheWAS meta-analyses adjusting for the effects of selected diagnostic statuses (i.e., tobacco use disorder) and related risk phenotypes (i.e., BMI). Up to 52 phecodes were excluded from these analyses due to the number of cases dropping below 100, resulting in 1468-1520 phecodes included in the conditional PheWAS. Overall, the patterns of associations remained largely consistent (**eFigure 2.1-2.6**, available at [*https://github.com/Yingjie1101/cross-disorder*](https://github.com/Yingjie1101/cross-disorder); **eTable 7.1-7.6**). After full adjustment, standardized regression coefficients compared to the original model changed by 6.6%-37.1% for associations with physical health conditions, and 13.6%-25.4% for associations with mental health conditions (see **eFigure 3** for OR distribution by disease category). Specifically, controlling for psychiatric diagnostic status reduced the standardized regression coefficients on average by 15.7%-27.4% (16.2%-28.7% for factor-specific diagnoses) across all medical conditions for each factor; thought and internalizing factors were affected most by this adjustment. Adjusting for tobacco use disorder further reduced the standardized regression coefficients by 1.7%-12.0%, with the most pronounced change concerning SUD-PGS. Additionally controlling for BMI resulted in minimal reduction in effect sizes (up to 2.3%). Despite these attenuations, the overall association patterns remained consistent. Interestingly, a number of PGS-disease associations become stronger after adjusting for psychiatric diagnostic status, including those for neoplasms, dermatologic and endocrine/metabolic conditions (**eFigure 4**), suggesting potential divergence between phenotypic comorbidity and underlying genetic risk.

#### **LabWAS: secondary trait–adjusted models**

As part of the sensitivity analyses, we sequentially controlled for diagnostic statuses and related risk phenotypes including tobacco use disorder and BMI on top of the baseline covariates (**eFigure 6.1-6.6, 7**). Overall, adjusting for all covariates reduced effect size estimates by 1.9%-34.9% compared to the original model. Controlling for diagnostic status reduced the effect on average only by 4%, while controlling for BMI reduced the magnitude of effects by an average of 15.4%. In the fully adjusted model, 15/27 unique measures remained significantly associated with at least one factor-PGS, with the majority remain metabolic markers (e.g., HDL.C, triglyceride, glucose, blood urea nitrogen, aspartate), immune markers (e.g., WBC, complement C4, CRP), and hematological markers (e.g, RDW, PCV).

**eFigure 1. Multivariate genetic structures of 13 major psychiatric disorders.** Standardized results from the correlated factor model (upper panel) and the hierarchical factor model (lower panel). AN, anorexia nervosa; OCD, obsessive-compulsive disorder; TS, Tourette syndrome; SCZ, schizophrenia; BIP, bipolar disorder; ASD, autism spectrum disorder; ADHD, attention-deficit/hyperactivity disorder; PTSD, posttraumatic stress disorder; MDD, major depressive disorder; ANX, anxiety disorders; AUD, alcohol use disorder; CUD, cannabis use disorder; OUD, opioid use disorder.
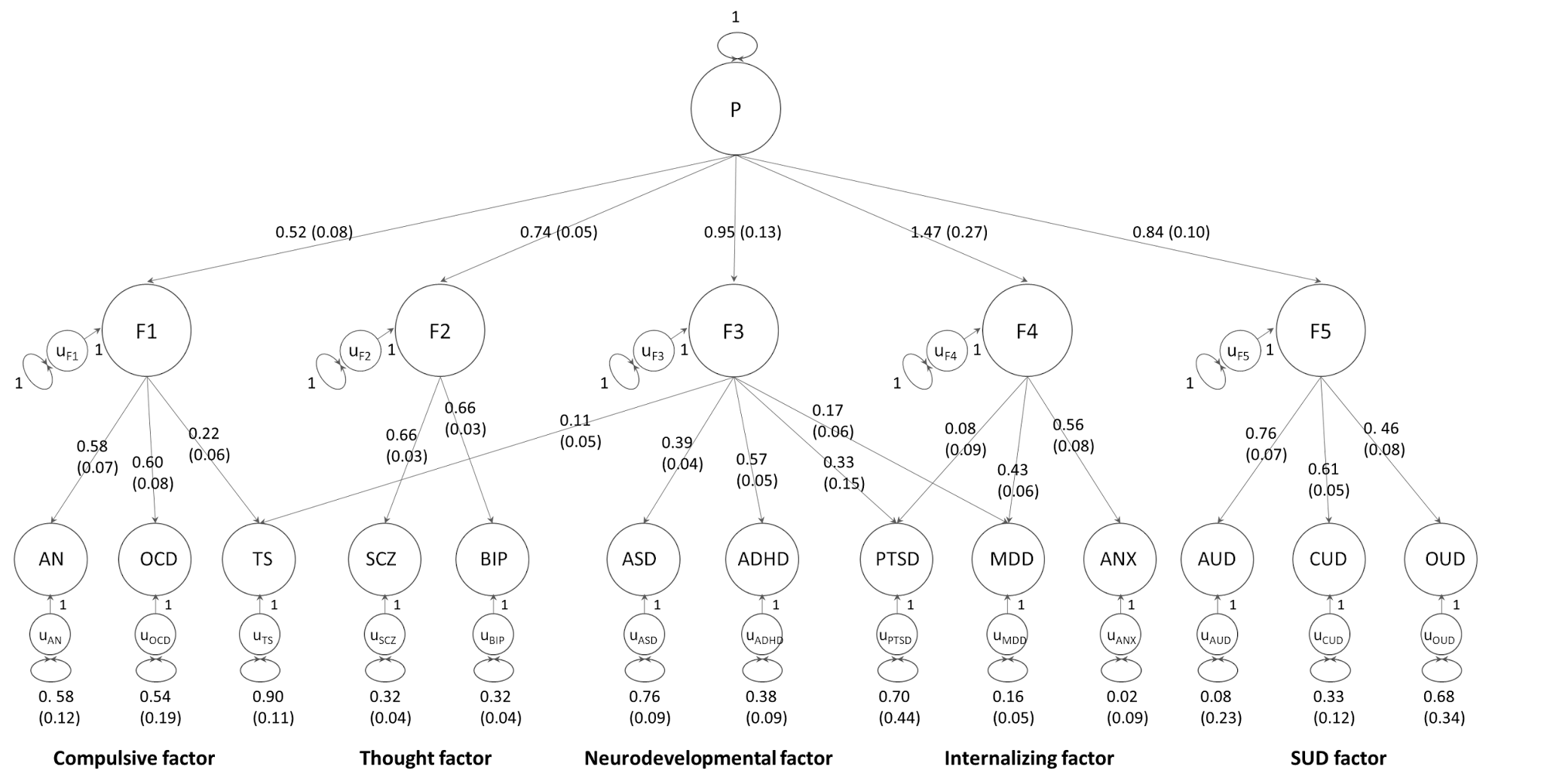

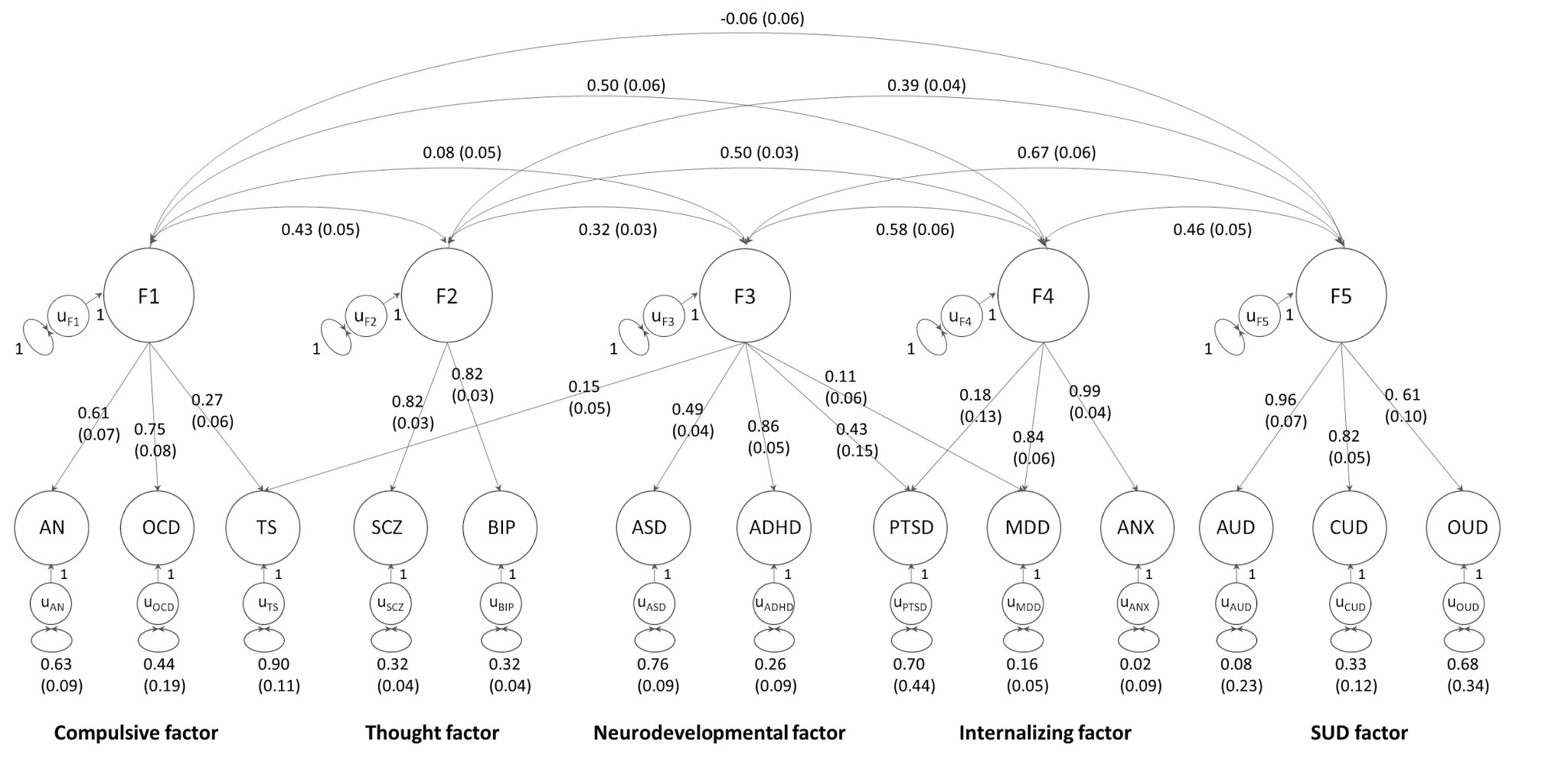

### **eFigure 2. Meta-analyzed primary and step-wise conditional PheWAS results.** Interactive Manhattan plots showing PheWAS Meta-analysis results in BioVU and MGBB, with one primary and four conditional PheWAS models. Each triangle represents association with a single medical phenotype (a total of 1,520 overlapping across both sites), with disease categories shown on the x-axis and -log10(p) shown on the y-axis. The direction of the triangles indicates the direction of effects. The grey solid line indicates Bonferroni-corrected significance threshold α=0.05/(1520*6)=5.48e-6 and the grey dotted line indicates α=0.05. Mouseover data points to see the corresponding phecode, beta and p values. Select areas to zoom in. Click on reset axes to return to the original figure. Subplots: cov0: primary PheWAS model adjusting for baseline covariates; Psyc-specific: additionally controlling for factor-specific diagnoses; Psyc-general: additionally controlling for factor-general diagnoses; Psyc+TUD: additionally controlling for factor-general diagnoses and Tobacco use disorder; Psyc+TUD+BMI: additionally controlling for factor-general diagnoses, Tobacco use disorder, and median body mass index.

[eFigure 2.1: Compulsive-PGS PheWAS](https://yingjie1101.github.io/cross-disorder/PheWAS_Comp.html)

[eFigure 2.2: Thought-PGS PheWAS](https://yingjie1101.github.io/cross-disorder/PheWAS_Thou.html)

[eFigure 2.3: Neurodevelopmental-PGS PheWAS](https://yingjie1101.github.io/cross-disorder/PheWAS_Neur.html)

[eFigure 2.4: Internalizing-PGS PheWAS](https://yingjie1101.github.io/cross-disorder/PheWAS_Inte.html)

[eFigure 2.5: SUD-PGS PheWAS](https://yingjie1101.github.io/cross-disorder/PheWAS_SUD.html)

[eFigure 2.6: Hierarchical-PGS PheWAS](https://yingjie1101.github.io/cross-disorder/PheWAS_Hier.html)

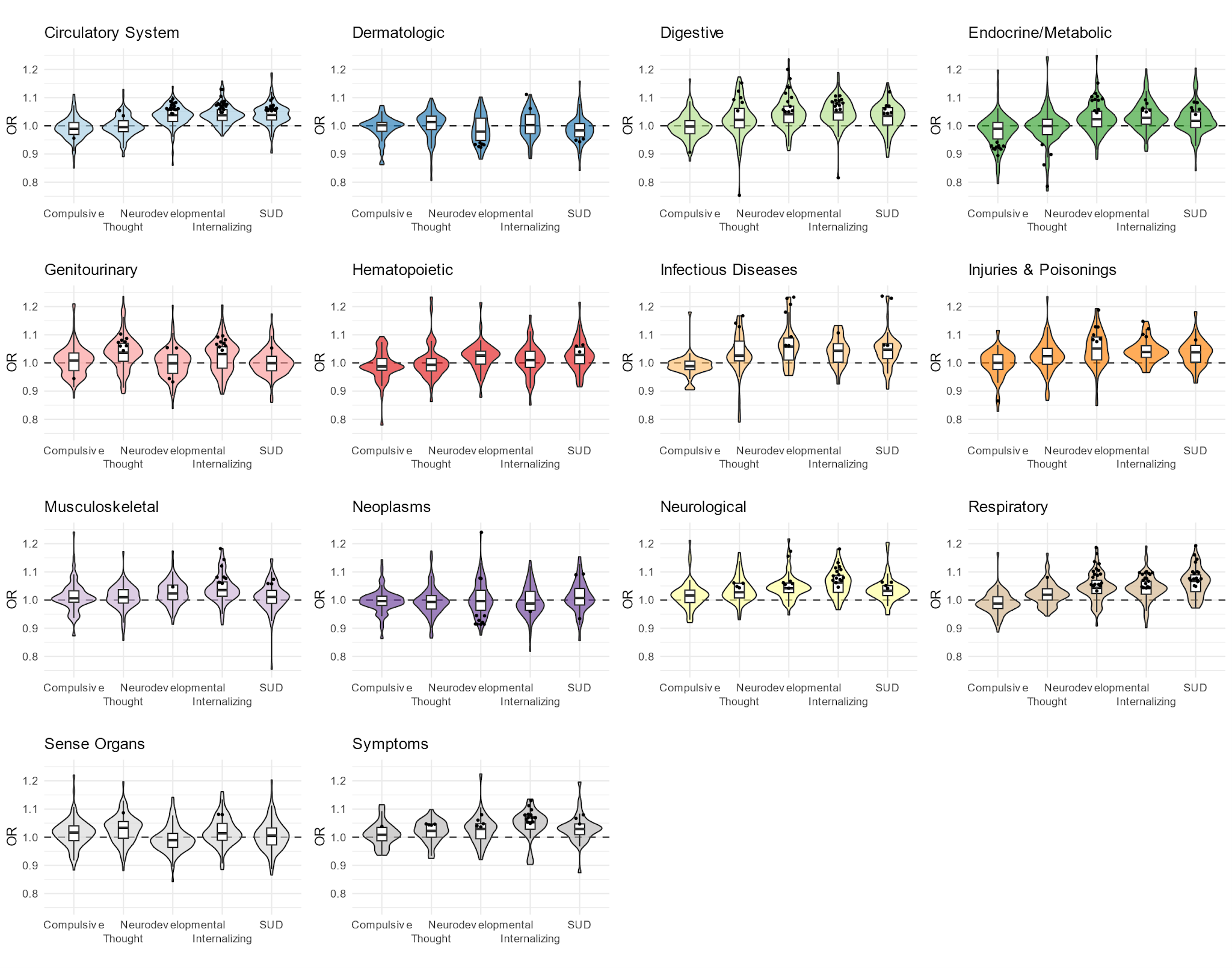

**Figure 3. The distribution of odds ratios (OR) for somatic disease categories across genomic factor PGSs.** Each sub-panel corresponds to a specific disease category, with the width of the violin plots indicating the distribution density of OR values, and boxplots show the interquartile range of ORs. Dots within the plots indicate significant associations with specific phecodes. Categories of congenital anomalies and pregnancy complications were excluded due to their lack of significant associations with any PGSs. SUD: substance use disorders.

### **eFigure 3. The distribution of odds ratios (OR) for somatic disease categories across genomic factor PGSs after full model adjustment.** Each sub-panel corresponds to a specific disease category, with the width of the violin plots indicating the distribution density of OR values and boxplots show the interquartile range of ORs. Dots within the plots indicate significant associations with specific phecodes. Categories of congenital anomalies and pregnancy complications were excluded due to their lack of significant associations with any PGSs. SUD: substance use disorders.

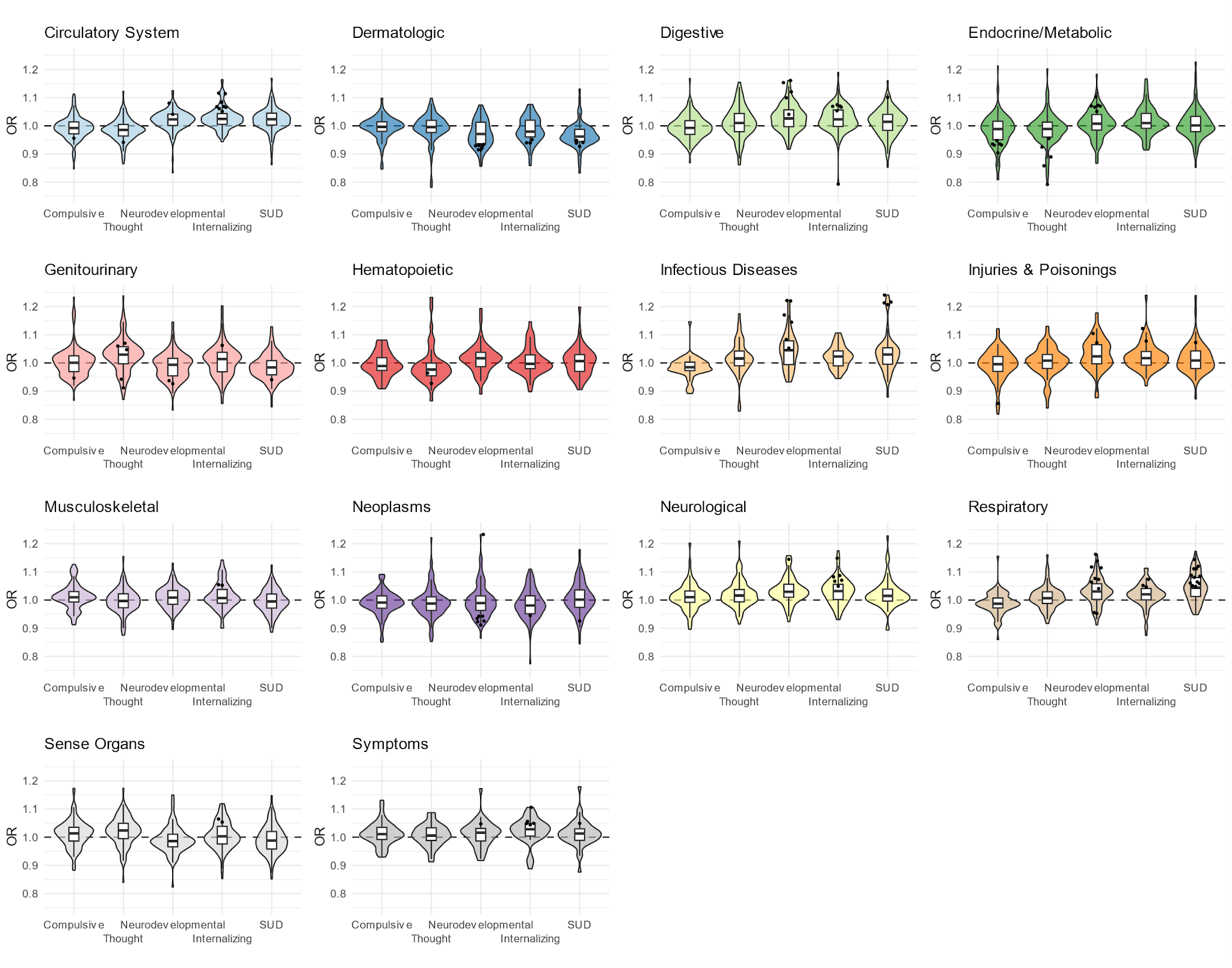

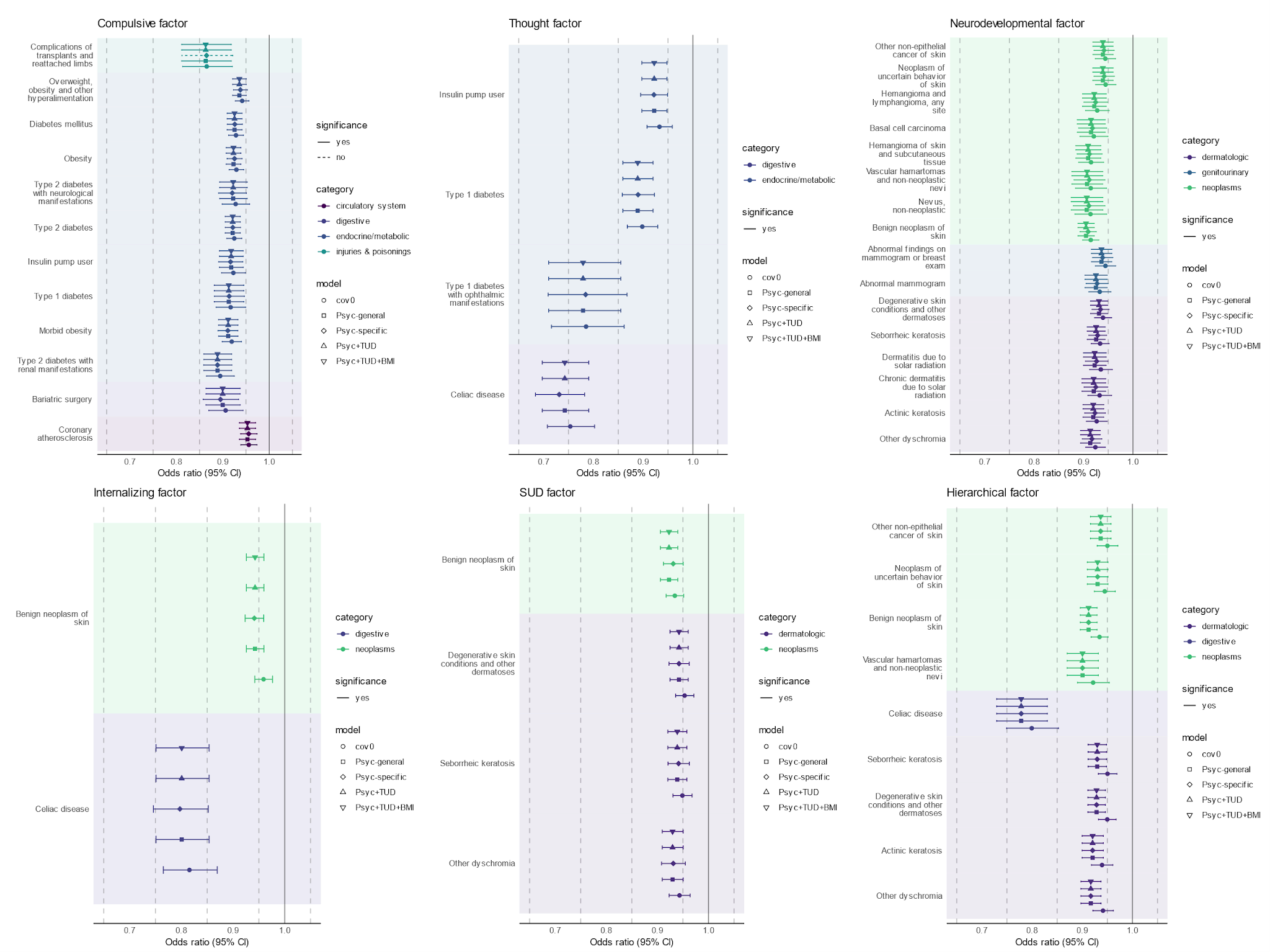

**eFigure 4. Top instances of associations with reversed effect size change in conditional PheWAS.** Among the significantly associated medical phenotypes for each genomic factor PGS, we presented the ones with the largest positive odds ratio change (stronger effect) in the four conditional PheWAS models, represented by different shapes of the dots. Color of the dots and background correspond to the somatic disease categories of the phenotypes. Error bar represents 95% confidence interval, with linetype represents statistical significance after Bonferroni correction. cov0: primary PheWAS model adjusting for baseline covariates; Psyc-specific: additionally controlling for factor-specific diagnoses; Psyc-general: additionally controlling for factor-general diagnoses; Psyc+TUD: additionally controlling for factor-general diagnoses and Tobacco use disorder; Psyc+TUD+BMI: additionally controlling for factor-general diagnoses, Tobacco use disorder, and median body mass index.

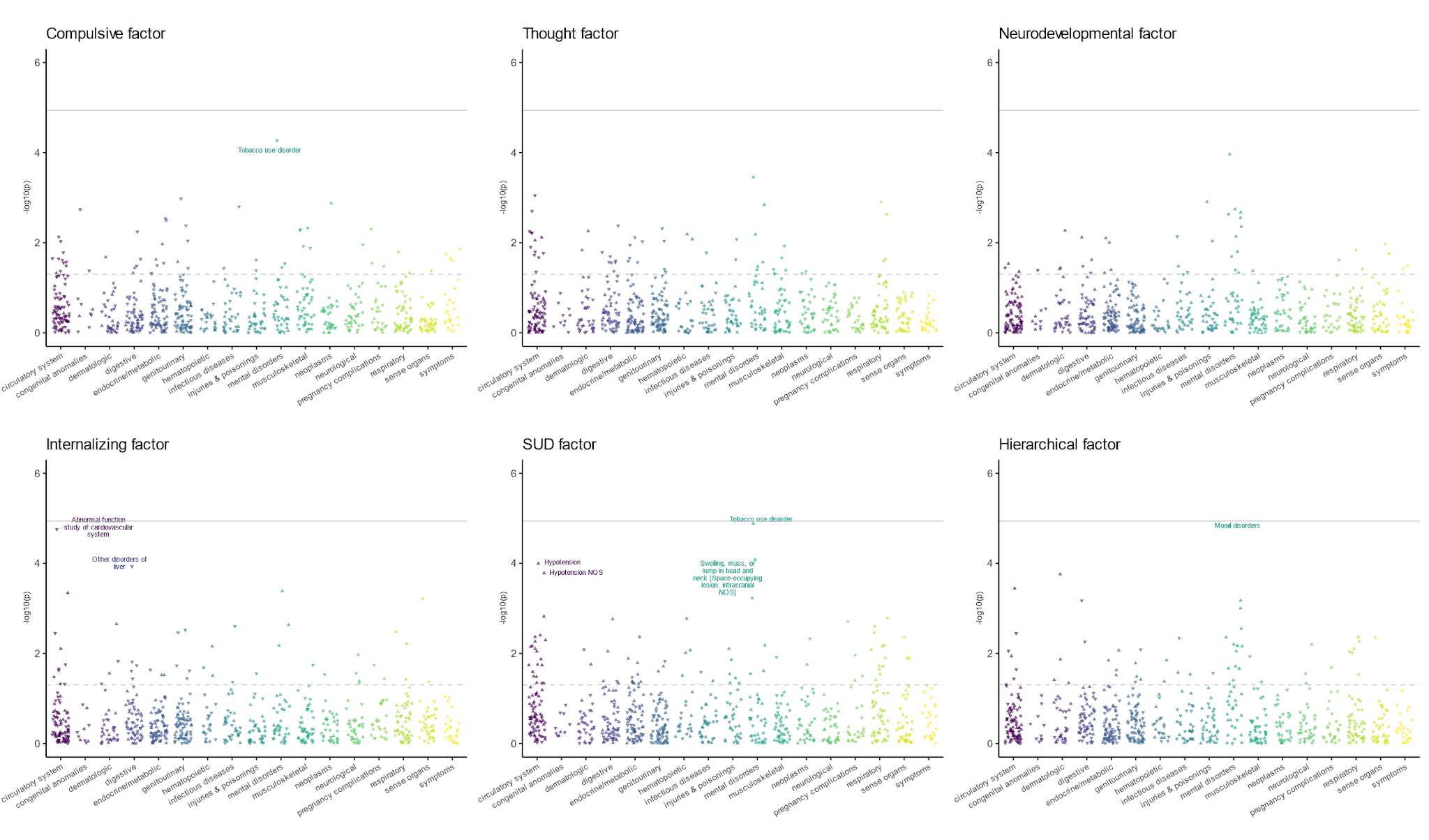

**eFigure 5. PheWAS of PGS for psychiatric genomic factors in BioVU African cohort.** We examined 724 disease phenotypes that had more than 100 cases available in this cohort (n=13,203). Phenotypes were grouped by disease category on the x-axis; the significance of each PGS-phecode association was shown on the y-axis as -log_10_(p). The direction of the triangle indicates the direction of effect. The grey solid line indicates Bonferroni-corrected significance threshold α=0.05/(724*6)=1.15e-5 and the grey dotted line indicates α=0.05. Significant associations that survived FDR correction were labeled. SUD: substance use disorders.

### **eFigure 6. Meta-analyzed primary and step-wise conditional LabWAS results.** Interactive Manhattan plots showing LabWAS Meta-analysis results in BioVU and MGBB, with one primary and four conditional LabWAS models. Each triangle represents association with a single medical phenotype (a total of 123 matched across both sites), with system categories shown on the x-axis and -log10(p) shown on the y-axis. The direction of the triangles indicates the direction of effects. The grey solid line indicates Bonferroni-corrected significance threshold α=0.05/(123*6)=6.78e-5 and the grey dotted line indicates α=0.05. Mouseover data points to see the corresponding phecode, beta and p values. Select areas to zoom in. Click on reset axes to return to the original figure. Subplots: cov0: primary LabWAS model adjusting for baseline covariates; Psyc-specific: additionally controlling for factor-specific diagnoses; Psyc-general: additionally controlling for factor-general diagnoses; Psyc+TUD: additionally controlling for factor-general diagnoses and Tobacco use disorder; Psyc+TUD+BMI: additionally controlling for factor-general diagnoses, Tobacco use disorder, and median body mass index.

### [eFigure 6.1: Compulsive-PGS LabWAS](https://yingjie1101.github.io/cross-disorder/LabWAS_Comp.html)

### [eFigure 6.2: Thought-PGS LabWAS](https://yingjie1101.github.io/cross-disorder/LabWAS_Thou.html)

### [eFigure 6.3: Neurodevelopmental-PGS LabWAS](https://yingjie1101.github.io/cross-disorder/LabWAS_Neur.html)

### [eFigure 6.4: Internalizing-PGS LabWAS](https://yingjie1101.github.io/cross-disorder/LabWAS_Inte.html)

### [eFigure 6.5: SUD-PGS LabWAS](https://yingjie1101.github.io/cross-disorder/LabWAS_SUD.html)

### [eFigure 6.6: Hierarchical-PGS LabWAS](https://yingjie1101.github.io/cross-disorder/LabWAS_Hier.html)

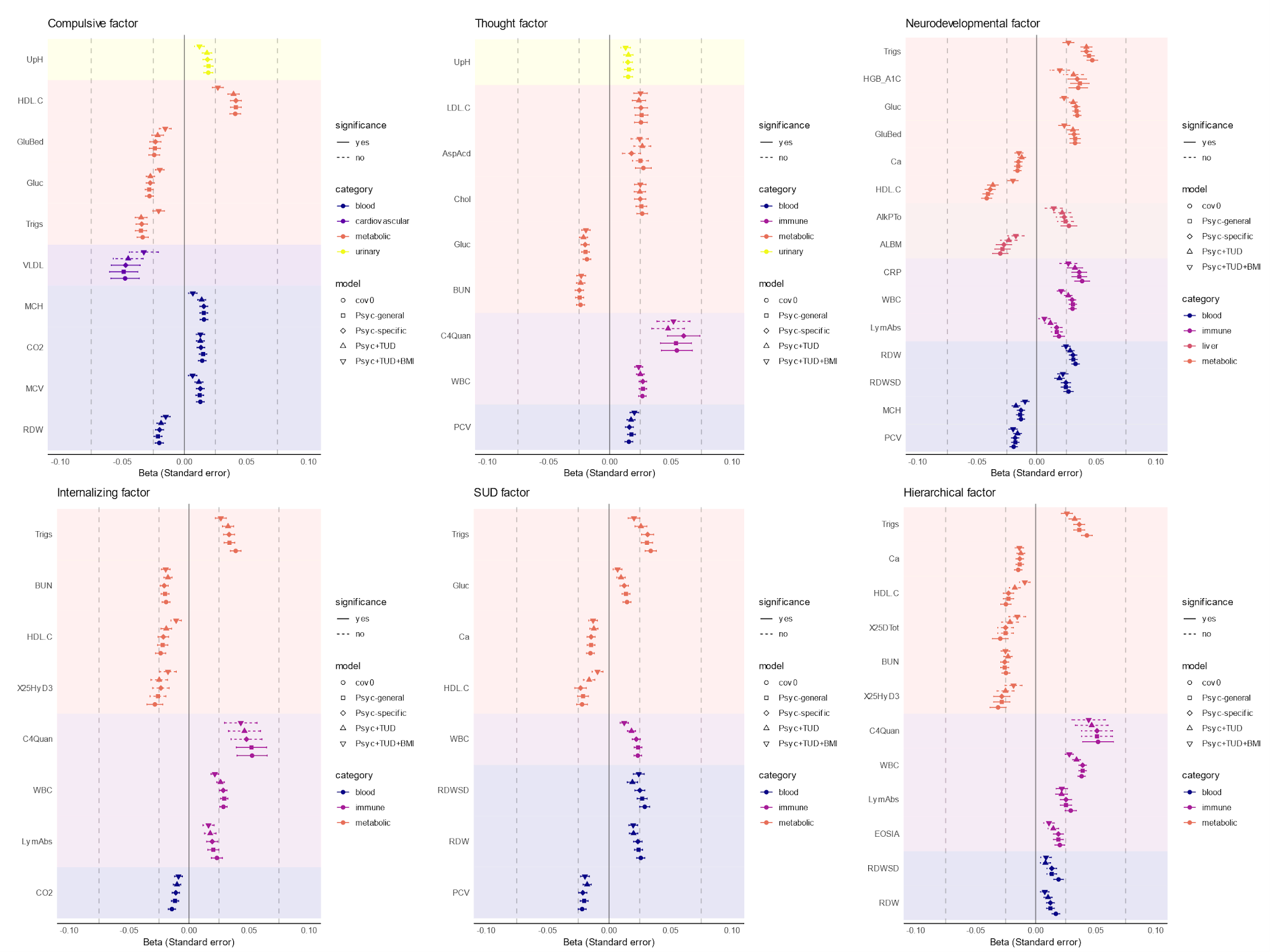

**eFigure 7. Effect sizes in primary and conditional LabWAS models.** For each genomic factor PGS, we selected labs that were significantly associated in the primary LabWAS model, and plotted their standardized beta values in the four conditional LabWAS models, represented by different shapes of the dots. Color of the dots and background correspond to the system categories of the labs. Error bar represents standard error, with linetype represents statistical significance after Bonferroni correction. cov0: primary LabWAS model adjusting for baseline covariates; Psyc-specific: additionally controlling for factor-specific diagnoses; Psyc-general: additionally controlling for factor-general diagnoses; Psyc+TUD: additionally controlling for factor-general diagnoses and Tobacco use disorder; Psyc+TUD+BMI: additionally controlling for factor-general diagnoses, Tobacco use disorder, and median body mass index.

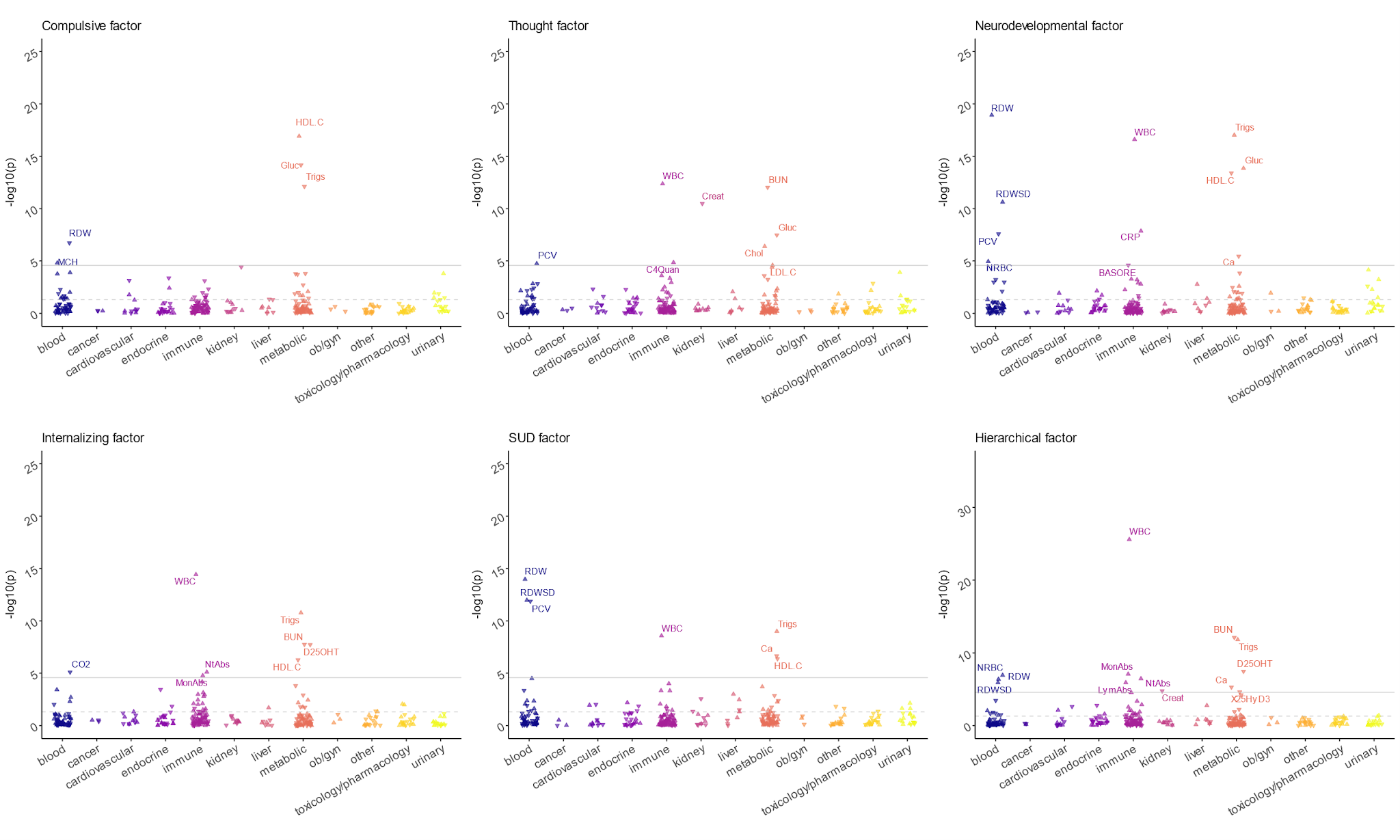

**eFigure 8. Laboratory-wide association scan (LabWAS) of PGS for psychiatric genomic factors at BioVU.** A total of 315 unique laboratory tests were included. Labs were grouped by system on the x-axis; the significance of each PGS-lab association was shown on the y-axis as -log_10_(p). The direction of the triangle indicates the direction of effect. The grey solid line indicates Bonferroni-corrected significance threshold α=0.05/(315*6)=2.65e-5 and the grey dotted line indicates α=0.05. Only significant associations are labeled. HDL.C, high-density lipoprotein cholesterol; Gluc, glucose; Trigs, triglyceride; RDW, erythrocyte distribution width [Ratio]; MCH, mean corpuscular hemoglobin; WBC, leukocyte (white blood cell) count; BUN, blood rea nitrogen; Creat, creatinine; Chol, cholesterol; C4Quan, complement C4; PCV, hematocrit; LDL.C, low-density lipoprotein cholesterol; RDWSD, erythrocyte distribution width; CRP, C reactive protein; Ca, calcium; NRBC, nucleated erythrocytes/100 leukocytes [Ratio]; BASORE, basophils/100 leukocytes; D25OHT,Vitamin D+Metabolites; NtAbs, neutrophil counts; CO2, carbon dioxide; MonAbs, monocyte counts; LymAbs, lymphocytes; X25HyD3, 25-hydroxyvitamin D3.

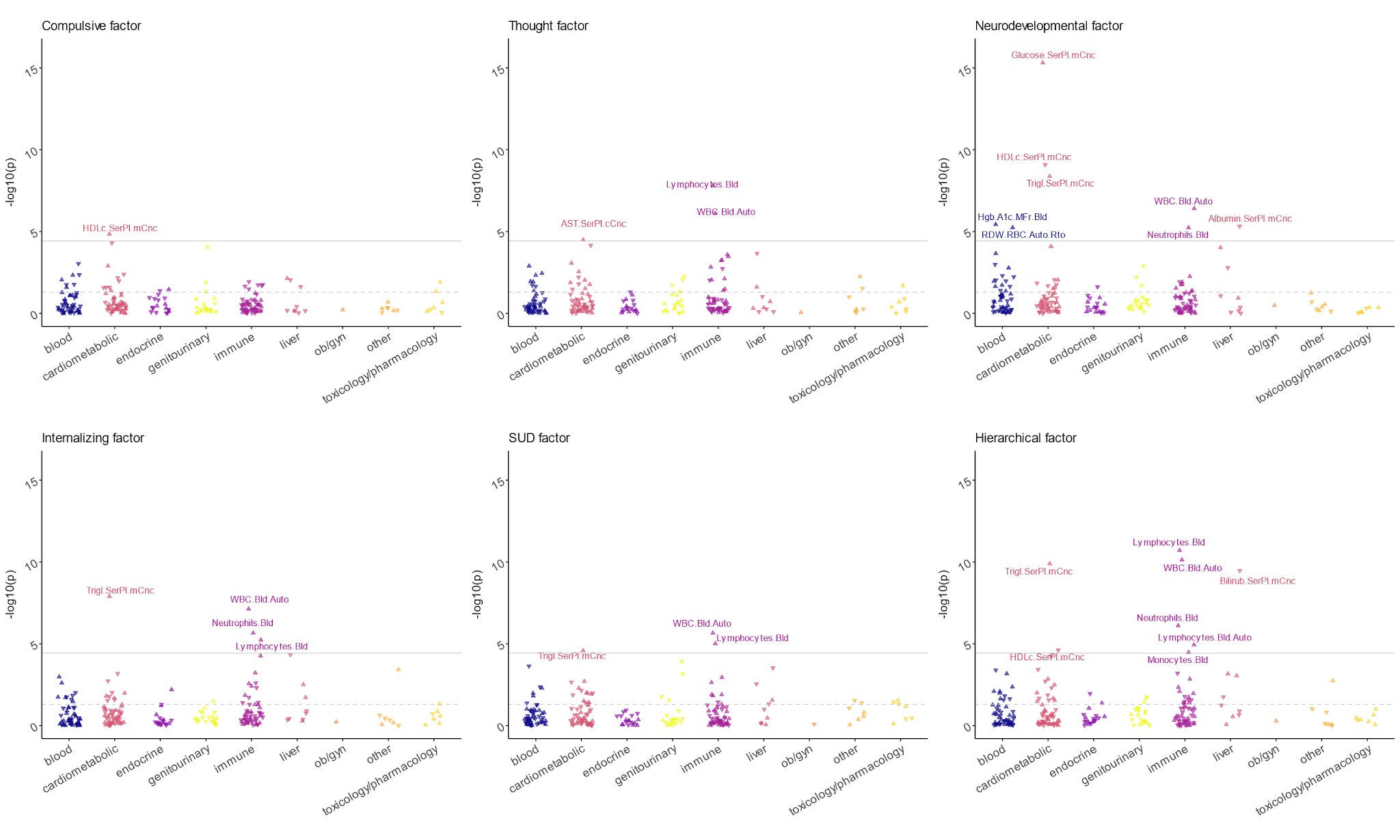

**eFigure 9. Laboratory-wide association scan (LabWAS) of PGS for psychiatric genomic factors at MGBB.** A total of 221 unique laboratory tests were included. Labs were grouped by system on the x-axis; the significance of each PGS-lab association was shown on the y-axis as -log_10_(p). The direction of the triangle indicates the direction of effect. The grey solid line indicates Bonferroni-corrected significance threshold α=0.05/(221*6)=3.77e-5 and the grey dotted line indicates α=0.05. Only significant associations are labeled. HDLc.SerPl.mCnc, high-density lipoprotein cholesterol; Lymphocytes.Bld, lymphocytes; WBC.Bld.Auto, white blood cells; AST.SerPl.cCnc, Aspartate aminotransferase; Glucose.SerPl.mCnc, glucose; Trigl.SerPl.mCnc, triglyceride; Hgb.A1c.MFr.Bld, Hemoglobin A1c/Hemoglobin total; Albumin.SerPl.mCnc, albumin; RDW.RBC.Auto.Rto, erythrocyte distribution width [Ratio]; Neutrophils.Bld, neutrophils; Bilirub.SerPl.mCnc, bilirubin total; Lymphocytes.Bld.Auto, lymphocytes; Monocytes.Bld, monocytes.

### **eReferences**

1.	Sealock, J. M. *et al.* Use of the PsycheMERGE Network to Investigate the Association Between Depression Polygenic Scores and White Blood Cell Count. *JAMA Psychiatry* **78**, 1365–1374 (2021).

2.	Dennis, J. K. *et al.* Clinical laboratory test-wide association scan of polygenic scores identifies biomarkers of complex disease. *Genome Med* **13**, 6 (2021).

3.	Grotzinger, A. D. *et al.* Transcriptome-Wide Structural Equation Modeling of 13 Major Psychiatric Disorders for Cross-Disorder Risk and Drug Repurposing. *JAMA Psychiatry* (2023) doi:10.1001/jamapsychiatry.2023.1808.

4. Ge, T., Chen, C.-Y., Ni, Y., Feng, Y.-C. A. & Smoller, J. W. Polygenic prediction via Bayesian regression and continuous shrinkage priors. *Nat Commun* 10, 1776 (2019).

5. Grotzinger, A. D. *et al.* Genomic structural equation modelling provides insights into the multivariate genetic architecture of complex traits. *Nat Hum Behav* **3**, 513–525 (2019).
